# Two Blood-based Endotypes Reveal Divergent Clinical Outcomes of Fibrotic Hypersensitivity Pneumonitis

**DOI:** 10.64898/2026.06.11.26355382

**Authors:** Yong Huang, Shwu-Fan Ma, John S. Kim, Emma Strickland, Brody A. Receveur, Catherine S. Bonham, Tessy K. Paul, Hannah C. Mannem, Numaan F. Malik, Jeffrey M. Sturek, Yun Michael Shim, Tania Velez, Samuel B. Konkol, In Su Cheon, Jie Sun, Ani Manichaikul, Ayodeji Adegunsoye, Mary E. Strek, Evan R. Fernández Pérez, Margaret L. Salisbury, Amy Zhao, Naftali Kaminski, Angela L. Linderholm, Manoj V. Maddali, Anne I. Sperling, Justin M. Oldham, Fernando J. Martinez, Imre Noth

**Author notes:** **Corresponding author** Imre Noth, MD, University of Virginia, Department of Medicine, Division of Pulmonary & Critical Care Medicine, PO Box 800546, Charlottesville VA, 22908-0546, m.

## Abstract

**Rationale:** Fibrotic hypersensitivity pneumonitis (fHP) is an antigen-driven, life-threatening interstitial lung disease characterized by heterogeneous radiologic features, clinical outcomes, and treatment responses.

**Objectives:** To identify blood-based fHP endotypes that inform mechanism, prognosis and therapeutic response.

**Methods:** We performed integrative analyses of multi-compartment transcriptomic data derived from whole blood, peripheral blood mononuclear cells, bronchoalveolar lavage, and surgical lung biopsies, alongside circulating plasma proteomics. Multiple clustering algorithms were cross-compared to ensure robustness and reproducibility of endotypes identification. Immune cell composition was inferred using bulk RNA-seq deconvolution and annotated with BAL single-cell RNA-seq. Pathway activities were characterized using Gene Set Enrichment Analysis. Transplant-free survival (TFS) was evaluated for endotype and corticosteroid exposure by Kaplan-Meier methods, with hazard ratios analyzed using multivariable Cox proportional hazards models.

**Results:** Two molecular endotypes, lymphocytic-associated (L-fHP) and non-lymphocytic-associated (N-fHP), were identified and validated. L-fHP showed enrichment of adaptive immune signaling and lymphocyte predominance, whereas N-fHP demonstrated myeloid-cell activation with neutrophil and macrophage predominance. Corticosteroid exposure was associated with worse TFS in L-fHP but not in N-fHP after adjusting for age, sex, and baseline pulmonary function. Compared to L-fHP, N-fHP had poorer baseline pulmonary function, faster 12-month FVC decline, and shorter TFS. N-fHP also exhibited elevated neutrophil-associated markers, including matrix metalloproteinase-9, across paired transcriptomic and proteomic datasets, supporting a neutrophil-driven, cross-compartment disease process.

**Conclusion:** Multi-omic, multi-compartment analysis identifies two reproducible fHP endotypes with distinct clinical outcomes and corticosteroid responses, supporting a precision medicine approach beyond current clinical and radiologic classification.

## Introduction

Hypersensitivity pneumonitis (HP) is an interstitial lung disease (ILD) resulting from immune responses to antigenic exposure (1). ILDs affect about 650,000 people and cause roughly 25,000 deaths annually, with HP accounting for ∼15% of cases (0.9 persons per 100,000) (2). Fibrotic HP can progress to end-stage lung disease, but with substantial inter-individual variability suggesting variation in disease pathobiology. The variable responses to both antigen abatement and immunosuppressive therapy underscore the need for deeper biological understanding to enable personalized disease classification and therapy (3, 4).

The immune response in HP patients is classically described as adaptive, involving both humoral and cellular mechanisms, and encompassing type III (immune complex-mediated) and type IV (delayed-type) hypersensitivity reactions (5). These responses lead to predominantly lymphocytic and granulomatous inflammation (6). While T cell-mediated alveolitis represents the typical pattern, patients with more fibrotic progression can exhibit increased lung neutrophilia (7). Neutrophilic inflammation has been associated with fibrotic remodeling and disease progression, whereas alterations in T cell function may exacerbate immune dysregulation (8–10). Use of bronchoalveolar lavage (BAL) lymphocytosis has been inconsistent in its use for diagnosis, particularly in fibrotic disease, where it serves as a supportive but non-definitive marker and underscores the presence of multiple immune-mediated processes involving cell types other than lymphocytes (11–13).

HP classification has evolved from acute, subacute, and chronic to a reliance on high-resolution computed tomography (HRCT) fibrotic patterns, classifying fibrotic (fHP) and non-fibrotic forms, with poorer outcomes in fHP (1). Machine learning approaches to clinical clustering have identified prognostic factors, including antigen type and exposure level, and comorbidities (14). Genetic variants and telomere shortening have also demonstrated prognostic relevance, and transcriptomic signatures from peripheral blood mononuclear cells (PBMCs) have been linked to disease progression (15). However, these approaches often rely on clinical or singular molecular traits, to drive endotype discovery, limiting their ability to capture the complexity of chronic, multifactorial disease.

Given the limitations of current clinicopathologic criteria, an ideal disease classification framework would incorporate molecular features and pathobiology to enable more accurate diagnosis, prognostic stratification, and therapeutic decision-making (16). Our goal was to delineate biologically distinct and clinically meaningful molecular endotypes that are reproducible across independent cohorts and in relevant tissues.

## Methods

### Study cohorts

Pulmonary Fibrosis Foundation (PFF) Patient Registry consisted of whole blood transcriptome and plasma proteome of fHP and IPF participants. University of California at Davis (UCD) and National Jewish Health (NJH) cohorts consisted of whole blood and peripheral blood mononuclear cells (PBMCs) transcriptome of fHP patients, respectively. Study-specific protocols were approved by institutional review boards at each participating site. All participants were diagnosed using established clinical guideline (1). See Online Supplement for additional details.

### PFF cohort

*Sample collection, RNA isolation, RNA-Seq library preparation and sequencing.* See Online Supplement for additional details.

### Sample clustering and dimensionality reduction

DDRTree, short for Discriminative Dimensionality Reduction via learning a Tree, provides an implementation of the framework of reversed graph embedding (RGE). It projects high-dimensional data into a reduced dimensional space, captures the intrinsic structure and relationships within the data, and constructs a principal tree for visualization (17).

Unsupervised machine-learning (ML) Consensus clustering approach was used to investigate the optimal endotypes within data structure (18). The parameters of 80% subsampling with 100 iterations and a range of k=2-6 clusters were used to evaluate the stability of classifications (19).

### Fibrotic HP classifier construction and machine-learning (ML) prediction of endotype

We employed the Recursive Feature Elimination (RFE) procedure in R/CRAN package ‘caret’ to identify the relevant classifier features (20). See Online Supplement for additional details.

### Immune cells deconvolution

We used R package ‘immunedeconv’ to quantify the fraction of the immune cell types from bulk RNA-sequencing data (21). See Online Supplement for additional details.

### Survival analysis of endotypes and corticosteroid exposure

Transplant-free survival (TFS) was defined as the time from blood sampling to lung transplantation or death, whichever occurred first. Patients without an event were censored at last clinical follow-up. Survival probabilities were estimated using the Kaplan-Meier (KM) method with log-rank test. Cox proportional hazards (PH) regression was used to calculate hazard ratio (HR) and Wald test p-value, with multivariable Cox-PH models being used to adjust for potential confounders. See Online Supplement for additional details.

## Results

### Transcriptomic profiling in independent cohorts and across different tissues consistently identifies two fHP endotypes with distinct immune cell composition

An overview of the multi-omics fHP cohorts across multiple tissues (Figure S1A) and flowchart of discovery and characterization of fHP endotypes (Figure S1B) are illustrated. Characteristics of fHP patients with blood transcriptome of the PFF, UCD, and NJH cohorts displayed only minor differences in mean age and disease severity as assessed by the forced vital capacity percentage predicted (FVC-pp) at enrollment (Table 1).

**Table 1.** Demographics and clinical traits of fHP blood RNA-seq cohorts.

| Clinical demographics | Pulmonary Fibrosis Foundation (PFF) | University of California-Davis (UCD) | National Jewish Hospital (NJH) |
| --- | --- | --- | --- |
| Sex (M/F, N) | 58/68 | 22/18 | 20/17 |
| Age (Mean±SD) | 66.3±9.2 | 71.0±9.4 | 62.2±10.4 |
| Race (Caucasian/Others, N) | 117/9 | 33/7 | NA |
| Smokers (Yes/No, N) | 56/66 | 17/23 | NA |
| <sup>1</sup>FVC %predicted (Mean±SD) | 66.2±20.0 | 61.5±20.5 | 72.5±13.1 |
| <sup>2</sup>D<sub>L</sub>CO %predicted (Mean±SD) | 44.1±15.7 | 43.2±17.9 | NA |
| CT <sup>3</sup>UIP (-/+, N) | 18/76 | NA | NA |
| <sup>4</sup>CS N; Days: IRQ (Range) | 39; 324.5 (11 - 2609) | NA | NA |
| <sup>5</sup>Immsup N; Days: IRQ (Range) | 34; 692 (2 - 1932) | NA | NA |
| <sup>6</sup>TFS (Dead/Alive, N) | 38/81 | 13/27 | NA |
| Follow-up month (Mean±SD) | 33.5±5.6 | 13.2±3.7 | NA |
| RNA-seq assay (N) | 126 | 40 | 37 |
| Proteomics assay (N) | 122<sup>#</sup> | NA | NA |

<sup>1</sup>FVC %predicted: Percent predicted forced vital capacity
<sup>2</sup>D<sub>L</sub>CO % predicted: Percent predicted diffusing capacity of the lungs for carbon monoxide
<sup>3</sup>UIP: Usual interstitial pneumonia
<sup>4</sup>CS: Corticosteroid exposure initiated before blood draw; Exposure Days before blood draw; Interquartile (IRQ)
<sup>5</sup>Immsup: Immunosuppressive agents; 21 patients overlap with CS
<sup>6</sup>TFS: Transplant-free survival
<sup>#</sup> Number of fHP patients in the PFF cohort with paired Olink™ proteomic data and bulk whole blood RNA-seq data
NA= Not Available

Machine-learning (ML) consensus K-means clustering of the PFF transcriptomic cohort samples identified k=2 as the most stable classification (Figure 1A), with a sharp block-diagonal structures in the consensus matrix, compared to k=3-6 (Figure 1B-C, Figure S2A-B). Cluster-consensus scores also demonstrated the highest stability in dichotomous classification (Figure S2C). Accordingly, we reduced the dimensionality of PFF transcriptome into two spaces using the Discriminative Dimensionality Reduction Tree (DDRTree) framework and confirmed the significant separation of the two groups by unsupervised Principal Component Analysis (PCA) (Figure 1D-E). We referred to the two groups as Minor and Major to reflect the sizes of the smaller and larger subgroups, respectively. Cross-comparison of ML Consensus clustering (k=2) with DDRTree bifurcation in the PFF cohort yielded 86.8% sensitivity (SE) and 100% specificity (SP) (Figure S2D).

**Figure 1.**
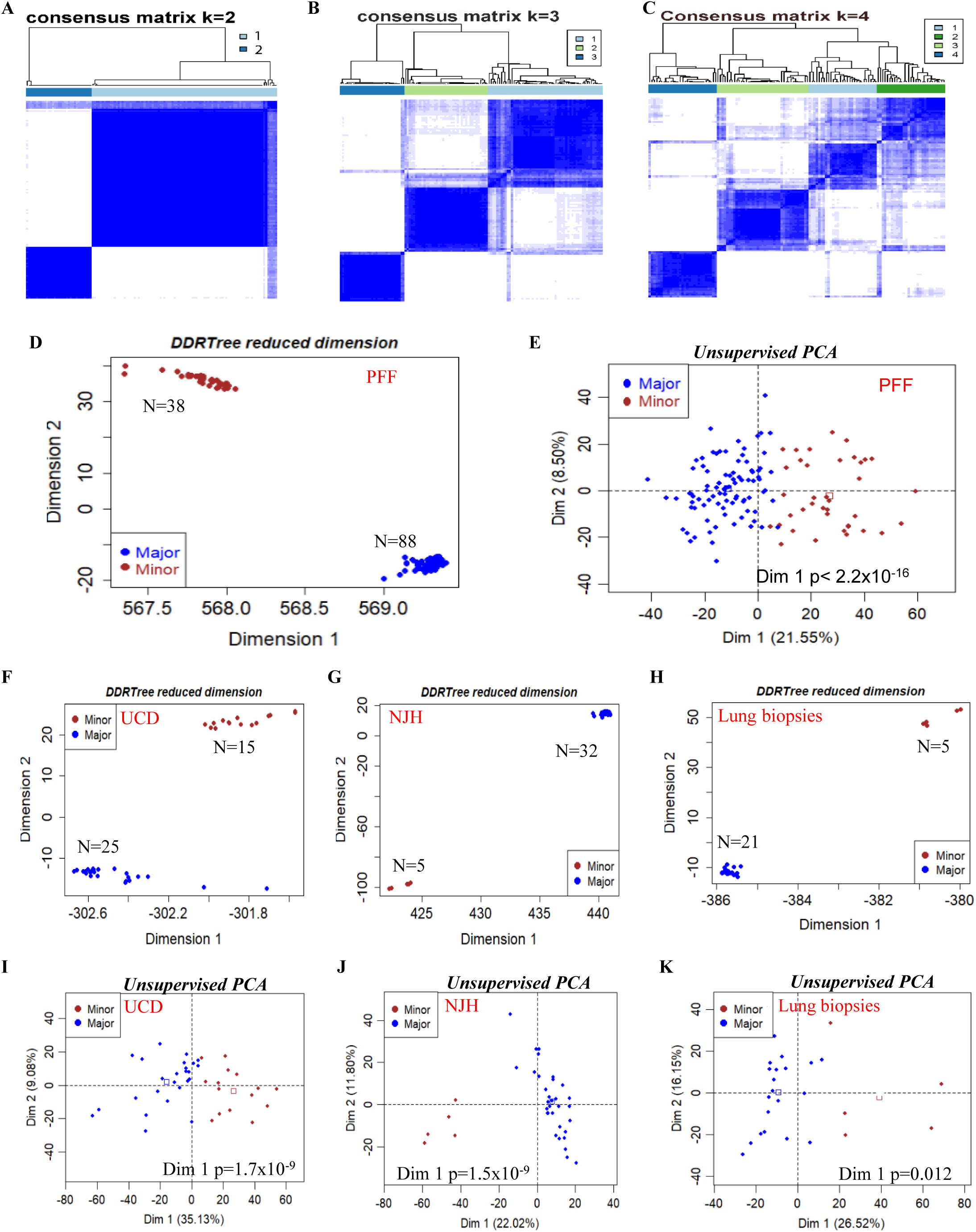
Endotype identification and validation across multiple blood cohorts and lung tissues using transcriptome data. (A-C) Unsupervised machine learning (ML) Consensus clustering of PFF with preset cluster k=2-4. The cluster distribution of 100 subsampling iterations displayed a clearer border when k=2, see Figure S2 for more details. (D) DDRTree dimension reduction reproducibly yielded a Major and a Minor class in PFF derivation cohort (see Figure S2A for cross-validation. (E) Unsupervised Principal Component Analysis (PCA) confirmed extremely significant separation of the Major and Minor class (p< 2.2×10^-16^). DDRTree dimension reduction yielded a Major and a Minor class in validation cohorts of (F) UCD whole blood, (G) NJH PBMC, and (H) surgical lung biopsies. Unsupervised PCA also verified Major and Minor class in (I) UCD whole blood, (J) NJH PBMC, and (K) surgical lung biopsies.

We repeated the DDRTree in each validation cohort with transcriptome data derived from whole blood, PBMCs, and lung surgical biopsies (GSE150910) (22) (Figure 1F-H; cohorts are described in Figure S1A). Unsupervised PCA of each cohort consistently showed a Minor and a Major group that was significant by two-group comparisons p-value for Principal Component 1 (Dim1) (Figure 1I-K).

Immune cell deconvolution revealed distinct immune cell profiles between two endotypes. The Minor group exhibited significant increase in neutrophil fraction, while the Major group exhibited significant increase in lymphocytes, specifically, B, NK and CD8+ T cells in whole blood transcriptome of PFF and UCD cohorts (Figure 2A-B, respectively). In NJH PBMCs transcriptome cohort without neutrophils, monocytes or CD8+ T cells were significantly higher in Minor or Major group, respectively (Figure 2C). The consistent profiles of immune cells in validation cohorts led us to designate the Major endotype as L-(Lymphocyte)-associated fHP (L-fHP) and the Minor endotype as N-(Non-lymphocyte)-associated fHP (N-fHP), respectively.

**Figure 2.**
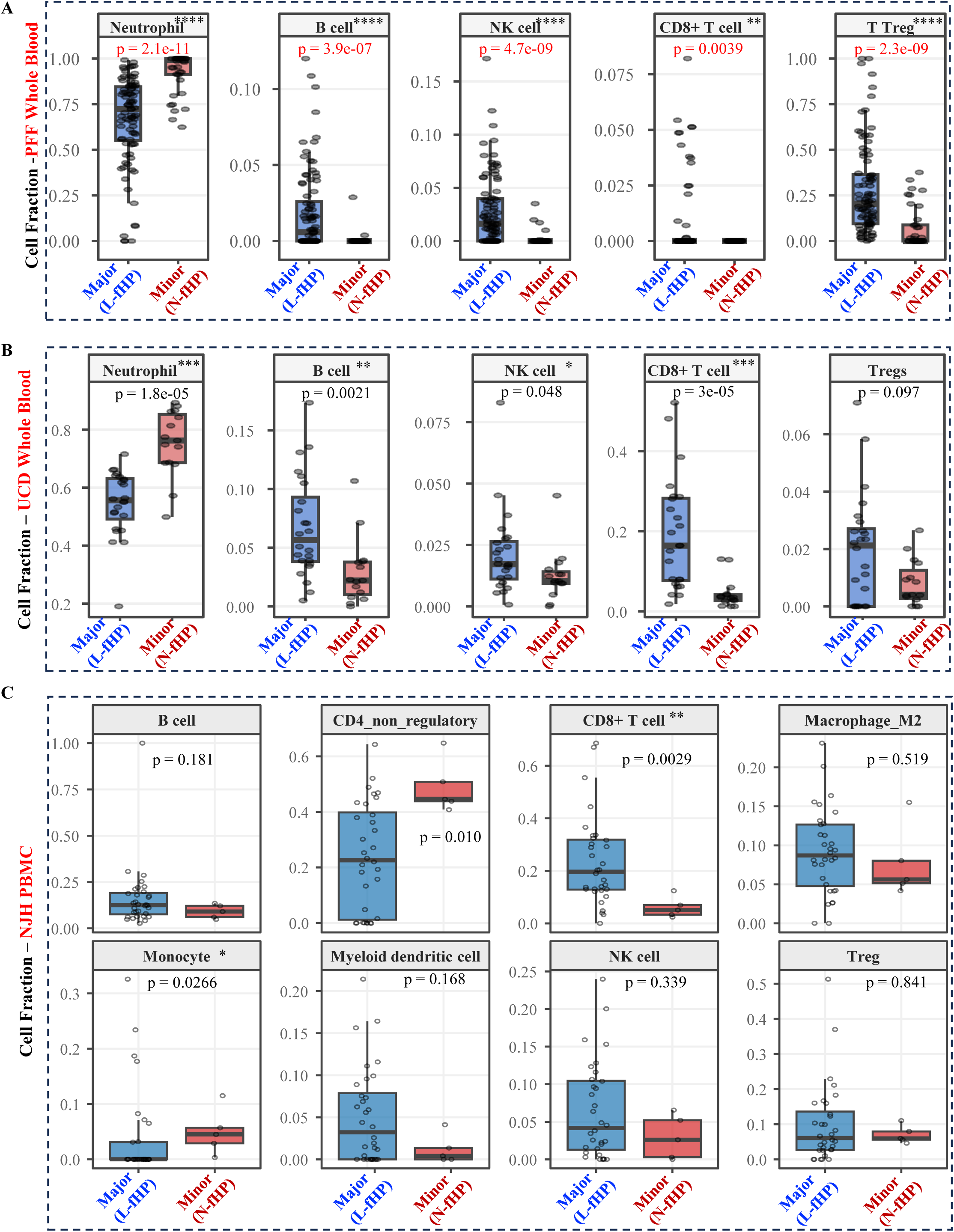
Immune cell deconvolution stratified by fHP endotypes. (A) PFF whole blood exhibited significantly increased fraction of neutrophil and decreased B cell, NK cell, CD8+ T cell and Treg cell in Minor compared to Major class. (B) UCD whole blood transcriptomic cohort exhibited significantly increased fraction of neutrophil and decreased B cell, NK cell and CD8+ T cell. (C) NJH PMBC exhibited significantly increased fraction of neutrophil and decreased CD8+ T cell in Minor compared to Major class. Accordingly, the Major class is designated as Lymphocyte-associated fHP (L-fHP) endotype, and the Minor class as non-lymphocyte-associated fHP (N-fHP) endotype, respectively.

### Functional characterization of the fHP endotypes reveal distinct adaptive versus innate immune programs

We performed a Recursive Feature Elimination with Random Forest model to construct a transcriptomic fHP-endotype classifier consisting of 40 genes (Table S1) using the PFF as training cohort. Imbalanced Random Forest model supervised by the classifier predicted fHP endotypes with 97.4% sensitivity and 96.6% specificity in the PFF cohort by 5-fold cross-validation, and 66.7% sensitivity and 96% specificity in the external validation of UCD cohort, revealing a performance that favored specificity over sensitivity (Table S2).

Volcano plots exhibited the differentially expressed genes (DEG) between the two endotypes in PFF (Figure 3A) and UCD (Figure 3B) RNA-seq data. Venn diagram demonstrated highly significant overlap of DEG between PFF and UCD RNA-seq data with the same direction of changes (one-tailed hypergeometric distribution p<10^-300^, Figure 3C; Tabel S3-4 for DEG in PFF and UCD RNA-seq datasets, respectively).

**Figure 3.**
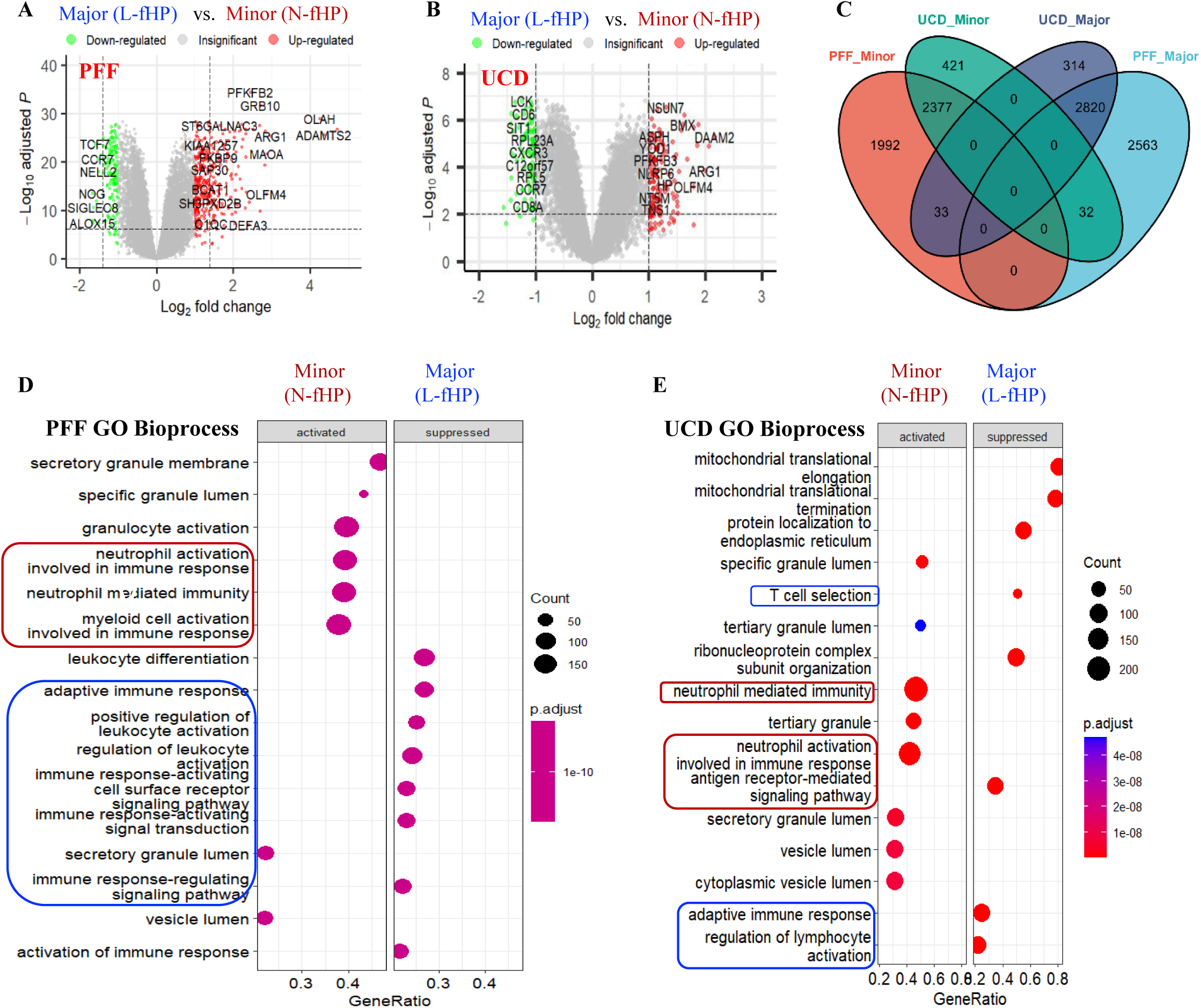
Functional characterization of fHP endotypes in the PFF and UCD cohorts. Volcano plots of differentially expressed genes (DEG) identified by eBayes moderated t-test between two fHP endotypes in (A) PFF and (B) UCD cohorts. Full DEG lists are displayed in Table S3-4. Red or green colors highlight genes which were up- or down-regulated with criteria of log2(fold change)>1 and FDR<0.05 in Minor (N-fHP) compared to Major (L-fHP), respectively. (C) Venn diagram of the full list of DEG revealed that 84% (2377/2830) of genes up-regulated in UCD Minor were also up-regulated in PFF Minor vs. Major. In concordance, 89% (2820/3167) of genes down-regulated in UCD Minor were also down-regulated in PFF Minor vs. Major. Gene Set Enrichment Analysis (GSEA) of Gene Ontology (GO) Bioprocess pathways of (D) PFF and (E) UCD cohorts revealed neutrophil and myeloid cell activation pathway in Minor/N-fHP (red box), and adaptive immune response pathway in Major/L-fHP group (blue box).

Insights into potential molecular mechanisms in each endotype were assessed using Gene Set Enrichment Analysis (GSEA). Gene Ontology biological process in both PFF and UCD cohorts again confirmed neutrophil and myeloid cell activation in N-fHP group, and adaptive immune response and leukocyte activation in L-fHP group (Figure 3D-E; see additional GSEA of PFF, UCD and NJH in Figure S3A-C, respectively, and lung tissues in Figure S4).

### Transcriptomic fHP-endotypes in the PFF cohort differ in baseline disease severity, longitudinal outcomes and corticosteroid (CS) response

At baseline, no difference was observed between N-fHP and L-fHP endotypes in sex, age, antigen exposure history, HRCT usual interstitial pneumonia (UIP) pattern, and corticosteroid or immunosuppressant usage prior to blood draw in the PFF cohort (Table 2; Figure S5). However, N-fHP patients consistently exhibited lower baseline forced vital capacity percent predicted (FVC-pp; p=0.002), lower baseline diffusing capacity of the lungs for carbon monoxide percent predicted (DLCO-pp; p=0.01), a more rapid decline in FVC-pp over 12 months (p=0.0037), and shorter TFS (p=0.001) (Table 2).These observations were replicated in the validation NJH and UCD cohorts significantly or by trend (Figure S6)

**Table 2.** Demographics and clinical traits of fHP endotypes identified in PFF cohort.

|  | N-fHP endotype | L-fHP endotype | *p*-val<sup>#</sup> |
| --- | --- | --- | --- |
| Sex (F/M, N) | 22/16 | 46/42 | 0.697 |
| Age (Mean ± SD ) | 67±9.2 | 65.9±9.3 | 0.53 |
| <sup>1</sup>Antigen type (B/D/F/M/O/U, N) | 13/3/3/3/4/11 | 20/4/1/20/8/35 | 0.066 |
| CT <sup>2</sup>UIP (+/-, N) | 5/23 | 13/53 | 1 |
| <sup>3</sup>Corticosteroid (+/-, N) | 16/22 | 23/65 | 0.32 |
| <sup>4</sup>IMS (+/-, N) | 11/27 | 23/65 | 0.91 |
| <sup>5</sup>FVC-pp (Mean ± SD) | 58.5±16.9 | 69.5±20.5 | 0.002* |
| <sup>6</sup>D<sub>L</sub>CO-pp (Mean ± SD) | 38.4±15.3 | 46.4±15.2 | 0.01* |
| FVC 12-month decline (Mean ± SD) | 12.8±8.0 | 2.4±7.0 | 0.0037* |
| <sup>7</sup>TFS (dead/alive, N) | 20/17 | 18/64 | 0.001* |

<sup>1</sup> Antigen type: Bird Fancier's lung/Down/farmer's lung/Mold/Other/Unknown; data is missed in one patient
<sup>2</sup> UIP: Usual interstitial pneumonia; CT data are unavailable in 32 fHP patients
<sup>3</sup> Corticosteroid: Corticosteroid treatment of fHP patients initiated before blood draw for RNA-seq
<sup>4</sup> IMS: Immunosuppressant treatment of fHP patients initiated before blood draw for RNA-seq
<sup>5</sup> FVC-pp: Percent predicted forced vital capacity
<sup>6</sup> D<sub>L</sub>CO-pp: Percent predicted diffusing capacity of the lungs for carbon monoxide
<sup>7</sup> TFS: Transplant-free survival at 36 month; Follow-up is lost in 7 fHP patients
<sup>#</sup> *p*-value: t-test for continuous data; Fisher's exact test for categorical data
\* significant *p* < 0.05

KM analysis revealed significantly different TFS across fHP endotypes and idiopathic pulmonary fibrosis (IPF) of the PFF cohort (global log-rank p=0.00062; Figure 4A). Pairwise Cox proportional hazards (PH) regression confirmed the increased hazard ratio (HR) of N-fHP endotype compared with L-fHP endotype or IPF (Figure 4B). In contrast, L-fHP exhibited improved TFS compared with N-fHP or IPF (Figure 4A-B). In a multivariable Cox-PH model adjusting for age, sex, FVC-pp, DLCO-pp and CS exposure, N-fHP patients still exhibited significantly worse TFS compared to L-fHP endotype patients, confirming the molecular endotype as a predictor of outcome (Figure 4C). Notably, CS exhibited no significant harm to the entire PFF cohort, underscoring the value of endotype stratification in prognostication (p=0.379; Figure 4C).

**Figure 4.**
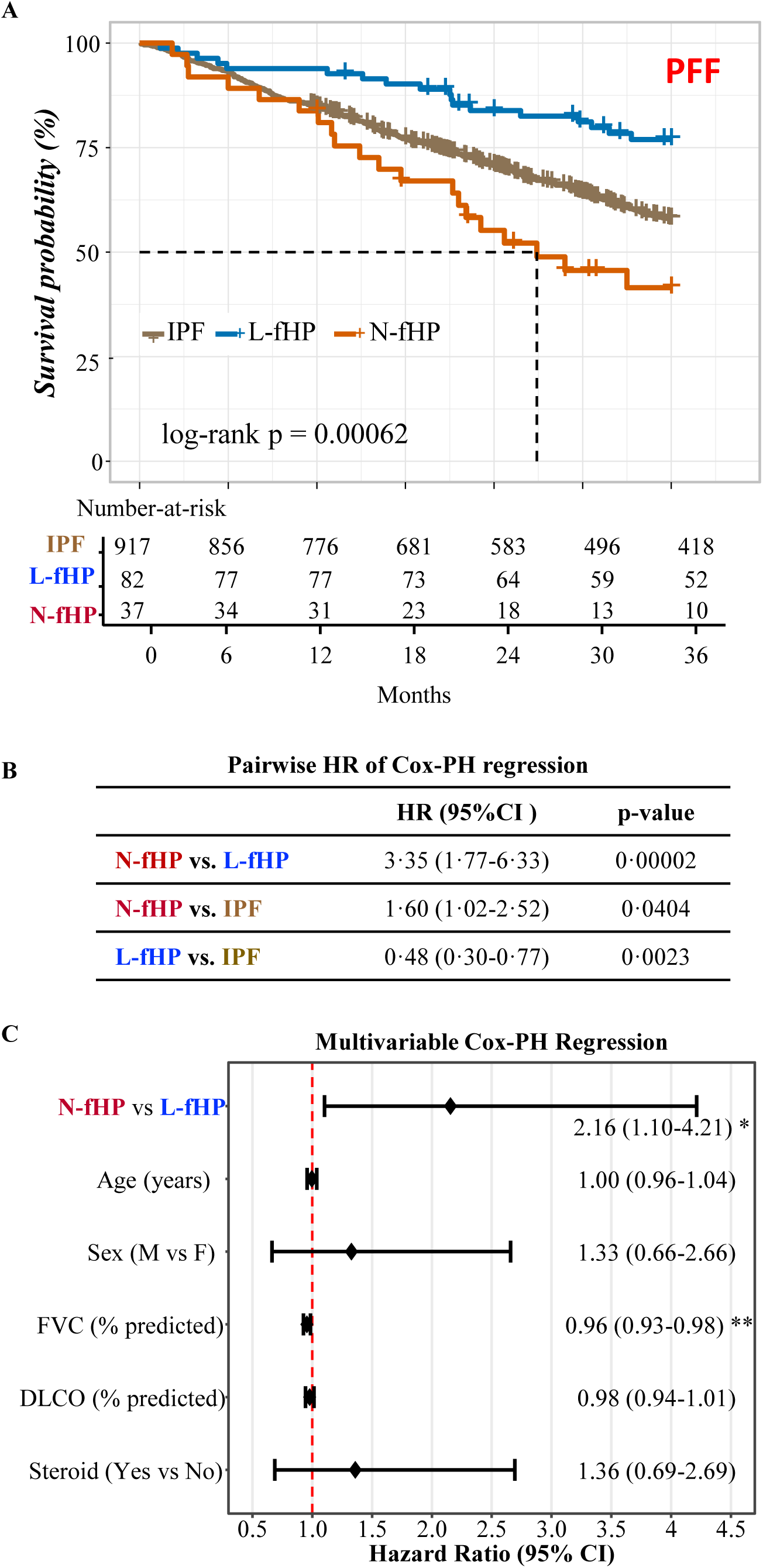
Association of fHP endotypes with transplant-free survival (TFS) in PFF cohort. (A) Kaplan-Meier (KM) plot of TFS analysis between N-fHP, L-fHP, and IPF of the PFF cohort revealed significant association with global log-rank *p*=0.00062. The rectangular dashed line denotes the critical time window where the survival curves show noticeable separation. (B) Cox-PH model demonstrated pairwise hazard ratios (HRs), 95% confidence intervals (95% CI) and Wald test *p*-values. (C) Forest plot of multivariable Cox-PH regression analysis of fHP endotypes adjusting for sex, age, FVC-pp, D_L_CO-pp and corticosteroid response. * or ** indicates p <0.05 or 0.005, respectively.

Examination of the impact of CS exposure that was initiated prior to blood sampling on survival outcomes surprisingly revealed significantly reduced TFS (log-rank p= 0.00014) and increased HR in L-fHP patients (HR=3.36, p=0.01), but not in N-fHP patients (HR=0.82, p=0.672) (Figure 5A-B). Multivariable Cox-PH model revealed a significant interaction between CS exposure and fHP endotypes (interaction HR=0.25, p=0.048; Figure 5C) and further confirmed endotype-specific CS response after adjustment of sex, age and baseline PFT (Figure 5C, CS p=0.036 in L-fHP, p=0.49 in N-fHP). Nevertheless, baseline FVC-pp (Figure S7A) and D_L_CO-pp (Figure S7B) did not differ between L-fHP patients with or without CS exposure.

**Figure 5.**
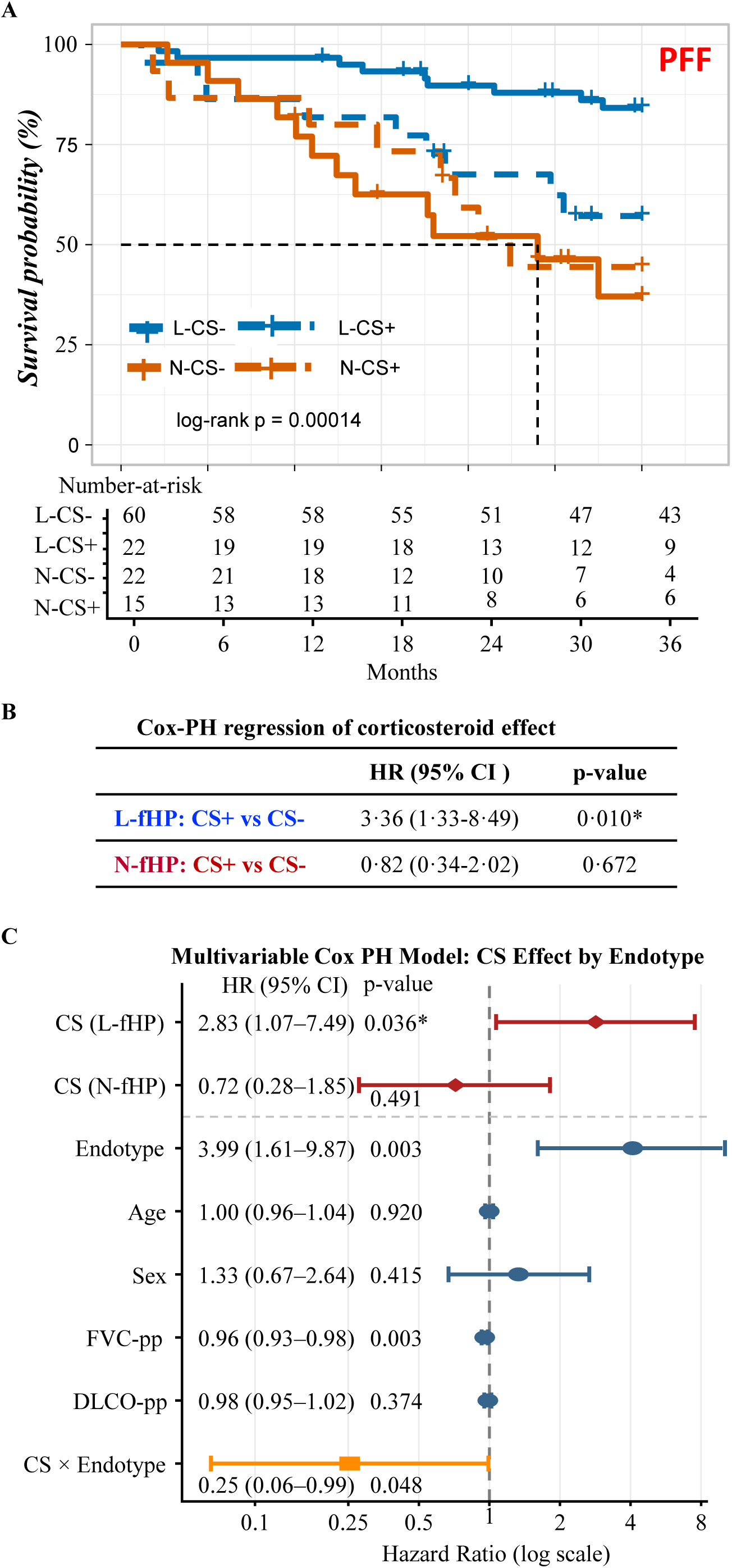
Association of fHP endotypes with corticosteroid (CS) response in the PFF cohort. (A) Kaplan-Meier (KM) analysis revealed significant reduction of TFS when treated by CS in L-fHP, but not in N-fHP patients (labeled as L-CS or N-CS, respectively). (B) CS exposure exhibited deleterious effect in L-fHP patients (HR=3.36, Wald p=0.01), but not in N-fHP patients. (C) Interaction analysis between CS exposure and fHP endotype (CS x Endotype) using a multivariable Cox-PH model revealed HR=0.25 and Wald p=0.048. Note that CS effect remained significant in L-fHP (p=0.036), but not in N-fHP (p=0.491), after adjusting for age, sex, FVC-pp, and D_L_CO-pp.

### BAL single-cell transcriptomics recapitulates blood transcriptome-defined fHP endotypes

A publicly available BAL scRNA-seq dataset (GSE271789) (23) with 9 HP patients enabled the conversion to pseudo-bulk RNA-seq and determination of two endotypes and their respective cell type compositions. Unsupervised ML consensus clustering again showed k=2 as the optimal cluster number (Figure S8A-G). DDRTree uncovered three samples in subclass-1 and six samples in subclasss-2 (Figure 4A) with 100% sensitivity and 100% specificity cross-compared to the ML Consensus clustering when k=2 (Figure S8H). Both methods confirmed that five out of six samples in Subclass-1 were fibrotic HP, while three samples in Subclass-2 were all non-fibrotic HP (Figure 6A). PCA supervised by the 40-gene fHP classifier (Table S1) further ensured significant separation of Subclass-1 and -2(p=0.016; Figure 6B). DEG between two subclusters are shown in Volcano plot (Figure 6C, Table S6).

**Figure 6.**
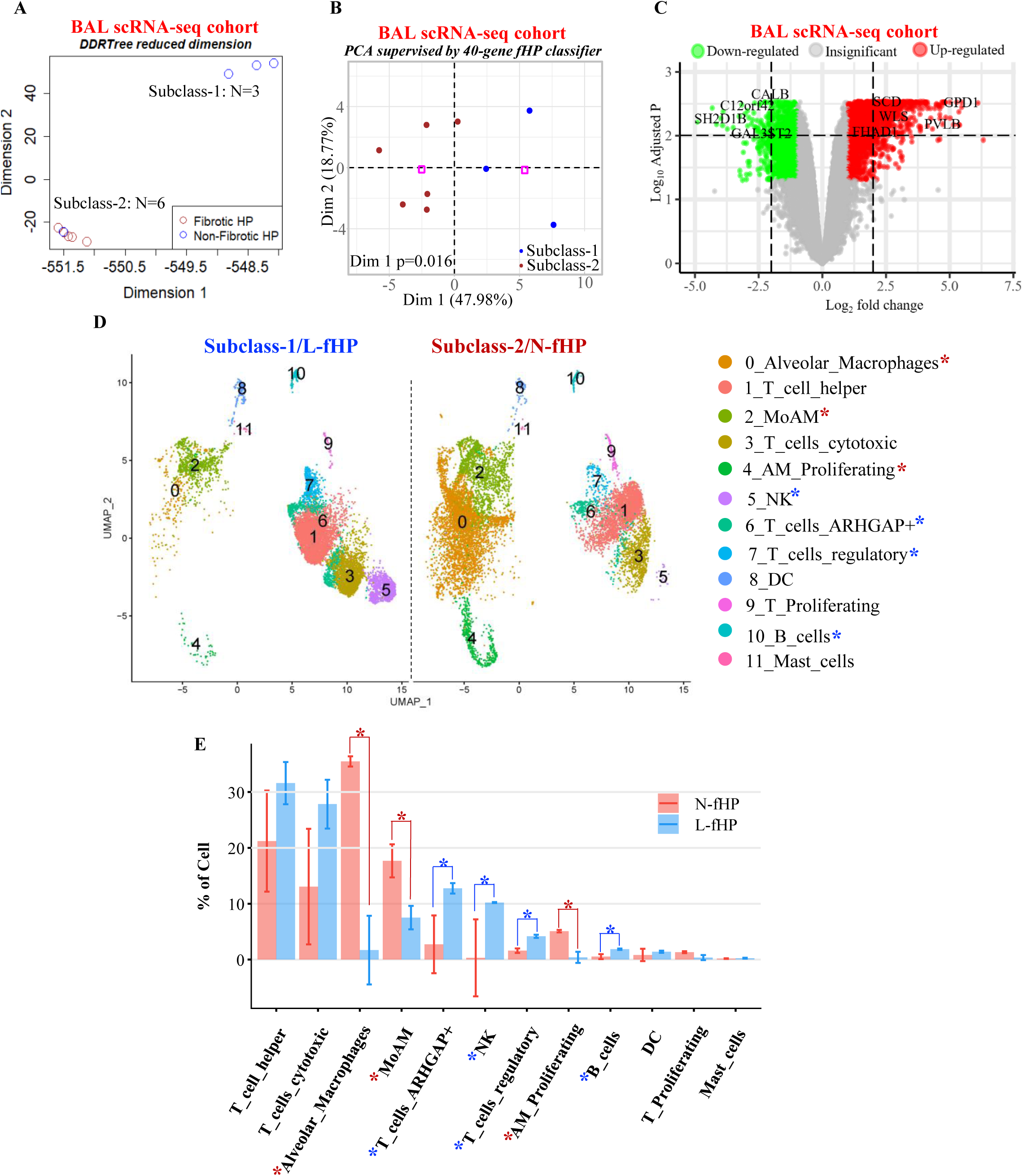
Endotype classification and cell type compositions in HP of bronchoalveolar lavage (BAL) scRNA-seq data (GSE271789). Pseudo-bulk RNA-seq analysis of GSE271789 BAL scRNA-seq data using (A) dimensionality reduction by DDRTree and (B) PCA supervised by fHP 40-gene classifier (see Table S1) yielded two reproducible subclasses. Subclass-1 consisted of 5 fibrotic HP and 1 non-fibrotic HP, while Subclass-2 consisted of 3 non-fibrotic HP. Cross-validation of DDRTree and machine-learning consensus clustering with k=2 exhibited 100% sensitivity and 100% specificity (see Figure S7.H). (C) Volcano plots of DEG identified by eBayes moderated t-test between two endotypes in PFF. Red or green colors highlight genes up- or down-regulated with criteria of log2(fold change)>1 and FDR<0.05 in Subclass-2 compared to Subclass-1, respectively. Pathway analysis based on DEG was displayed in Figure. S8. (D) UMAP of a random subset of 10,000 cells from each endotype of BAL scRNA-seq data. Asterisks indicate significantly increased cell types in L-fHP (blue) or N-fHP (red) endotype. (E) Statistic comparison of each cell type between two endotypes of the BAL 40555 cells in GSE271789. Asterisks represent significant difference between two endotypes (t-test p<0.05). Subclass-2 exhibited significantly higher fraction of macrophages (red asterisk in E), while Subclass-1 exhibited significantly enhanced fraction of T cells and B cells (blue asterisk in E). Therefore, Subclass-1 or 2 is assigned as L-fHP or N-fHP, respectively.

Uniform Manifold Approximation Projection (UMAP) of a random subset of 10k cells from L-fHP ad N-fHP illustrated changes of cell types with more T and B and NK cells in Subclass-1 (blue asterisks in Figure 6D), and more macrophages (MP) in Subclass-2 (red asterisks in Figure 6D). Statistical comparison of the cell fraction revealed significantly higher alveolar macrophages (Ave_MP), proliferating alveolar macrophages (AM_Proliferating) and monocyte-derived alveolar macrophages (MoAM) in Subclass-2 (red asterisks in Figure 6E), while higher proportion of T cells, Treg, and B cells in Subclass-1 endotype (blue asterisks in Figure 6E). Therefore, Subclass-1 and -2 were designated as L-fHP and N-fHP, respectively. GSEA of the endotypes in BAL pseudo-bulk RNA-seq data affirmed two immune subtypes like those observed in the whole blood transcriptome cohorts (Figure S9).

### Plasma proteomics confirms transcriptomic fHP-endotypes in the PFF cohort and provides insights into potential pathogenesis

We analyzed the circulating plasma Olink proteomics derived from the same patients for PFF RNA-seq and confirmed the immune subtypes of the two fHP endotypes. Specifically, we identified 213 significantly differentially expressed proteins (DEP) between the two transcriptomic endotypes (FDR<0.05, Figure 7A; Table S7). Again, GSEA of DEP confirmed neutrophil activation and myeloid leukocyte mediated immunity in N-fHP and adaptive immune response in L-fHP endotype (Figure 7B).

**Figure 7.**
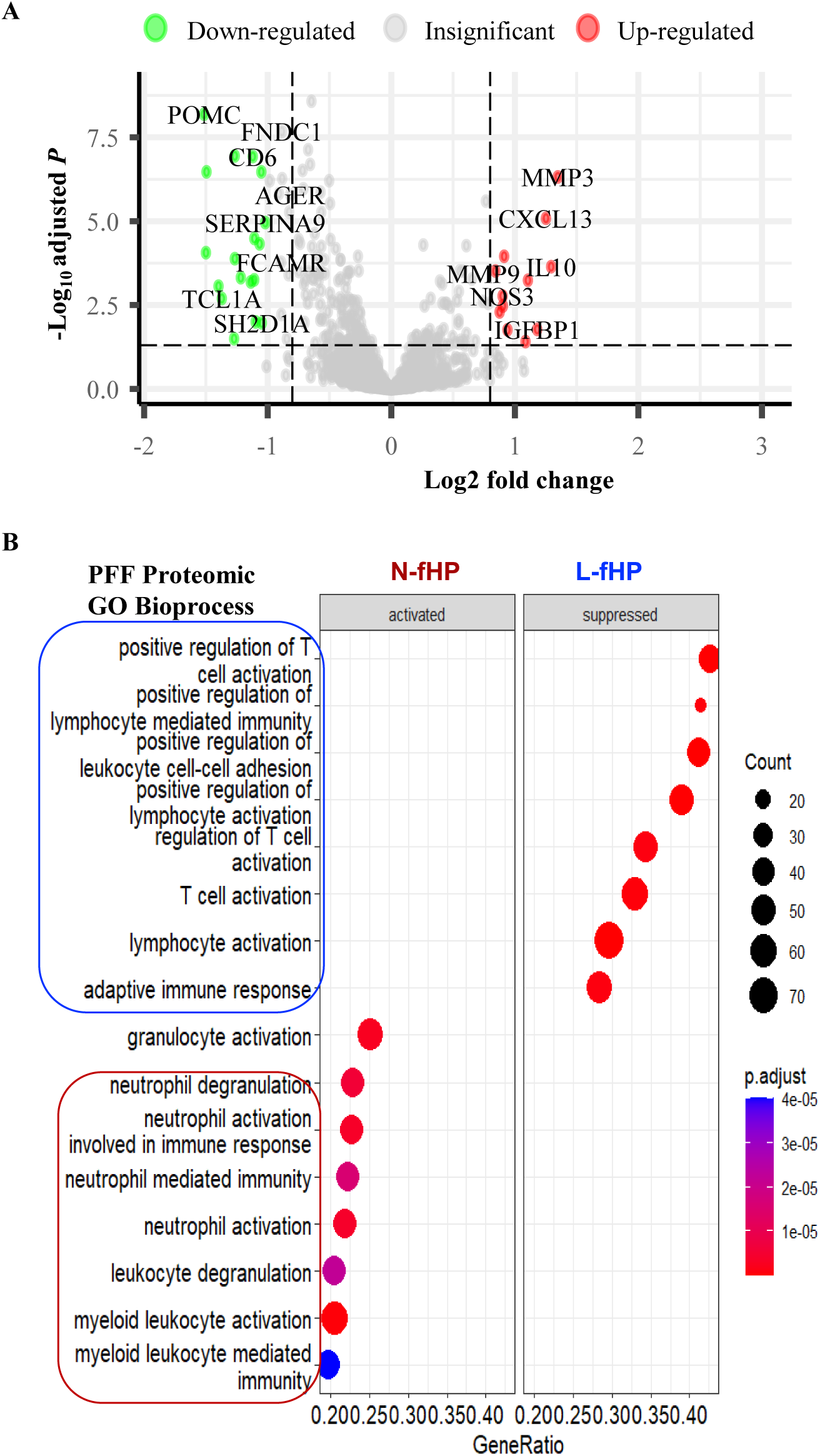
Validation of fHP endotypes by the circulating plasma proteomics in the PFF cohort. (A) Volcano plot of differentially expressed proteins (DEP) between fHP endotypes. Red and green dots represented significantly up- or down-regulated proteins in N-fHP compared to L-fHP, respectively. (B) GSEA for GO biological process revealed neutrophil activation and myeloid leukocyte mediated immunity in N-fHP (red box), and activated adaptive immune response mediated by lymphocytes or T cells in L-fHP (blue box).

Hierarchical clustering of neutrophil markers in PFF transcriptome revealed N-fHP associated genes involved in vascular basement membrane injury and neutrophil and monocyte recruitment (Figure S10). We further examined Matrix metalloproteinase-9 (MMP9) as a representative effector in paired proteomics and transcriptomics data derived from the same patients in PFF cohort. Notably, *MMP9* transcript expression was strongly increased in the N-fHP endotype, with near-complete penetrance (36/38 patients above the cohort median) and extreme significance in whole-blood transcriptome of PFF cohort (p<2.2×10^-16^, Figure 8A). Concordantly, MMP9 protein levels in N-fHP were also significantly higher than in N-fHP (p=0.007; Figure 8B). Indeed, MMP9 protein abundance was only significantly correlated with transcript level in N-fHP patients (p=0.022; Figure 8C) but not in L-fHP patients, indicating efficient transcript-to-protein coupling specific to this endotype. Consistent with a neutrophil-dependent mechanism, MMP9 and Myeloperoxidase (MPO) upregulation in N-fHP were detected exclusively in whole-blood transcriptome containing abundant neutrophils (Figures S11A and S12A-B, respectively), but not in any transcriptome lacking neutrophils (PBMCs, surgical lung biopsies, and BAL; Figure S11B-D, S12C).

**Figure 8.**
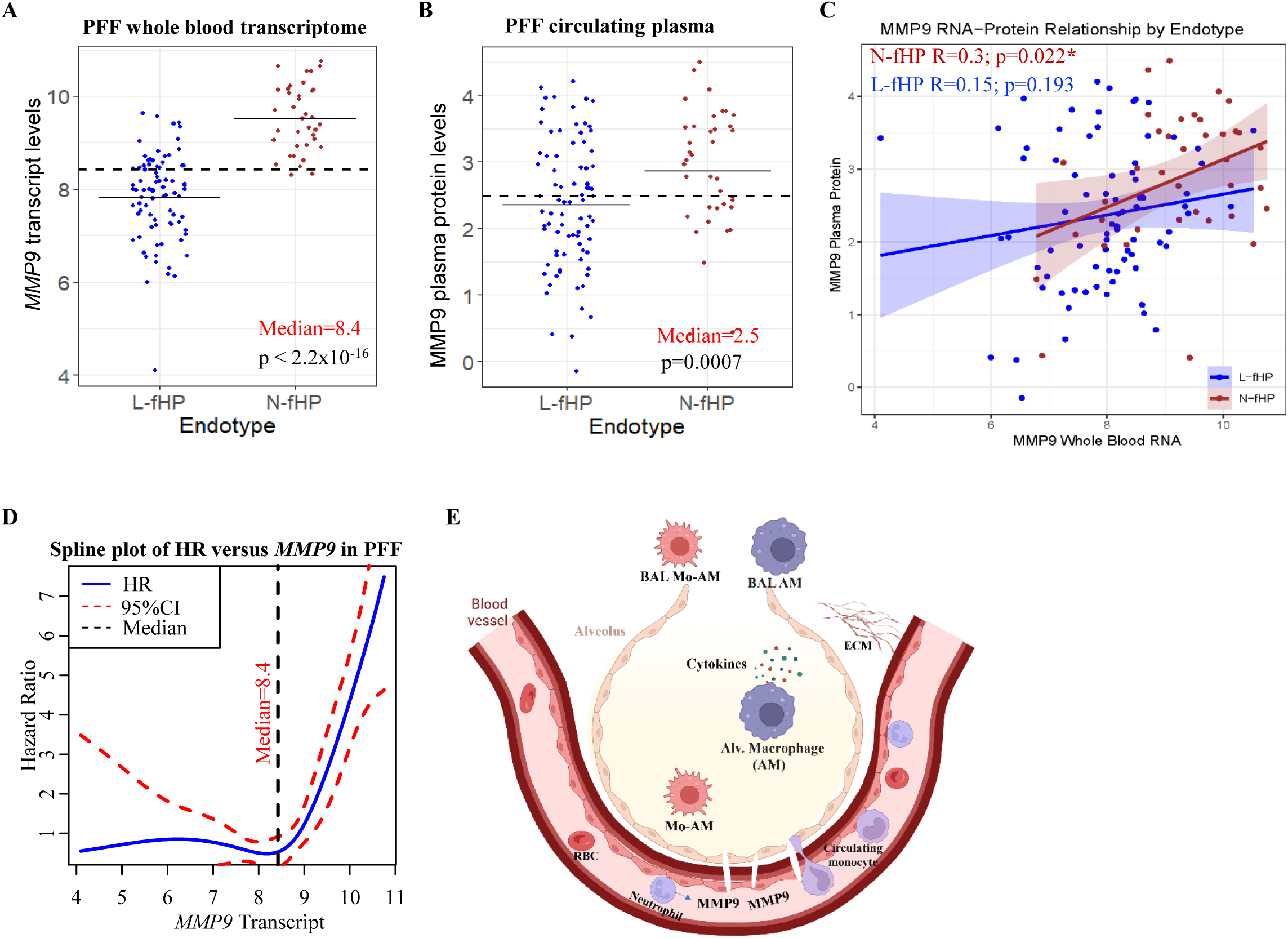
Paired whole blood transcriptome and circulating plasma proteome revealed potential mechanism of N-fHP. (A) Neutrophil marker Matrix metalloproteinase-9 (MMP9) transcript levels in whole blood of RNA-seq and (B) protein levels in circulating plasma of Olink proteomics were significantly higher in N-fHP than L-fHP of the PFF cohort. Solid line represents the endotype-specific average, while dashed line represents cohort median. (C) Endotype-stratified correlation of *MMP9* transcript and protein levels in paired whole blood cells and plasma derived from the same patients in PFF. Correlation was significant in N-fHP (p=0.022), but not in L-fHP (p=0.193). (D) Spline plot of hazard ratio of TFS against the *MMP9* gene expressions in whole blood transcriptome demonstrated a dose-response relationship once values were above the average level of MMP9 protein. Dash lines represent the overall median level of MMP9. (E) Schematic diagram of hypothesized tissue interactions in N-fHP pathogenesis: (1) Neutrophil activation and release of MMP9; (2) Breakdown of pulmonary vascular basement membrane by MMP9; (3) Migration of circulating monocytes into alveoli; (4) Alveolar macrophage activation and release of cytokines; (5) ECM and collagen deposition.

Functionally, spline regression of TFS revealed a non-linear association of hazard ratio with whole blood MMP9 transcript, with drastically increasing HR emerging once MMP9 expression exceeded the cohort median (Figure 8D). Together, these findings support a model in which peripheral blood neutrophil-released enzymes act as a cross-tissue effector driving fibrotic remodeling in lung and adverse outcomes in the N-fHP endotype, as summarized in the schematic diagram (Figure 8E).

## DISCUSSION

Our findings support two molecular endotypes of fHP with divergent immune responses, validated across independent cohorts and tissues. Importantly, these endotypes associate with different clinical outcomes and corticosteroid treatment responses, likely dependent upon distinct underlying immune processes. Our study is the first to use an unbiased approach, relying solely on cross-tissue multi-omics data, to identify fHP endotypes, in contrast to supervised methods that use clinical traits, comorbidities, or disease progression (14, 24, 25).

Lymphocyte cellular analysis has historically been a central focus in fHP, particularly in BAL, where it has been used as a diagnostic guide (1). However, this emphasis on adaptive immune signatures may have obscured the contribution of innate immune activities. Using whole blood transcriptomics, we identified a non-lymphocytic-associated (N-fHP) endotype characterized by enrichment of neutrophil-associated pathways and myeloid immune activation, supported by paired circulating plasma proteomic endotyping. Neutrophilic inflammation is recognized to play a role in both early disease and subsequent fibrosis, primarily based on only a few studies that have demonstrated neutrophil recruitment in lung tissue, associated with IFN-γ production in the acute phase and granuloma formation in HP development (7, 10, 26). N-fHP of BAL exhibits a significant increase of monocyte-derived alveolar macrophages (MoAMs), which are implicated in fibrosis, and ECM deposition in lung biopsies (27). Persistent macrophage activation may play a critical role in the long-term tissue destruction and fibrotic changes observed in fHP.(28, 29) Furusawa *et al.* noted that activation of epithelial cell pathways in HP lung is associated with poorer lung function.(22) In concordance, the N-fHP endotype demonstrates increased ciliated epithelial activity in the NJH PBMCs transcriptomic cohort (30).

Analysis of available BAL scRNA-seq data also supports two endotypes, with L-fHP enriched in lymphocytosis samples and N-fHP enriched in non-lymphocytosis samples. Current differential diagnosis of fHP and non-fHP is based on lymphocyte cellular analysis of BAL and transbronchial or surgical lung biopsy. BAL lymphocytosis greater than 30% has been advocated to separate chronic HP from IPF in the setting of a UIP pattern (1). Blood-based molecular endotyping represents a scalable and minimally invasive approach to differential diagnosis of HP. Importantly, transcriptomic endotypes of BAL align well with those of lung biopsies where N-fHP is characterized by increased collagen-containing ECM activity. BAL fluid assays often reveal variable lymphocyte counts (31). A higher percentage of BAL lymphocytosis has been associated with better long-term survival,(7, 11) and has been a feature of HP that potentially used to drive therapeutic selection (32). The long-standing paradigm in HP has been one of immunological dysregulation consisting of both humoral and T-helper cell type 1 (Th1) cellular immune responses that lead to a lymphocytic inflammatory pattern (1, 33). The argument has been that prominent BAL lymphocytosis is a defining element (13). Our BAL findings showing higher percentages of T cells identified by scRNA-seq, confirmed the expectation of the L-fHP endotype and demonstrated that BAL lymphocytosis portends a better prognosis, in accord with prior literature (11, 32). We would argue that the literature to date, including the BAL lymphocytosis studies, have been only describing this larger of the two endotypes, L-fHP.

With higher FVC-pp relative to N-fHP, the L-fHP appears to represent somewhat milder disease phenotype with more favorable outcomes than IPF, which is congruent with published data on HP in general (34, 35). This lymphocyte-associated profile is also consistent with an analysis of local lung tissue transcriptome from fHP patients which identified six molecular traits, in which regions with less extensive fibrosis exhibited the strongest T-cell activation pathways (26). Similarly, Furusawa *et al*. have also reported that adaptive immune–dominant gene expression profiles were associated with better lung function in fHP (22). Our findings suggest that the lymphocytic endotype may underlie these previously observed associations.

The impact of fHP endotype on clinical outcomes and response to therapy is striking, if surprising. While N-fHP endotype FVC-pp was lower at baseline, longitudinal analysis support that being a result of a more rapid 12-month FVC-pp decline, rather a lead time bias. TFS was best in L-fHP patients, followed by IPF, and worst in N-fHP. Yet, the two endotypes displayed no differences in age, sex, or HRCT UIP pattern. N-fHP remained significantly associated with worse TFS than L-fHP after adjustment for demographic variables, baseline disease severity, and CS exposure before blood draw, further assuring the prognostic utility of molecular endotypes, particularly in guiding early referral for lung transplantation.

The observed interaction between CS exposure and fHP endotype provides evidence that these endotypes reflect biologically and clinically distinct disease states. Systemic exposure to CS or immunosuppressive agents starting before blood sampling, and radiologic UIP pattern did not differ by endotype. CS impact with worsened TFS of L-fHP remained significant after adjustment of demographic variables and baseline pulmonary function. In contrast, no adverse effect of CS exposure on TFS was observed in N-fHP patients, suggesting that the deleterious impact of CS on outcome may be solely attributable to adaptive immune status rather than to innate immunity. Indeed, prior studies also reported an association between CS exposure with reduced survival in fHP, albeit non-significant (36, 37). Taken together, the use of CS as standard therapy in fHP warrants re-evaluation, with consideration given to molecular endotypes and immunologic heterogeneity when guiding treatment decisions (36, 37).

Our proteomics paired to transcriptomics data in the PFF cohort enabled several insights that provide additional biological validation of the existence of two endotypes. The neutrophil-derived matrix metalloproteinase MMP9, also known as gelatinase B, emerged as a representative effector of the N-fHP endotype in current study. Whole-blood MMP9 transcript levels were markedly increased in N-fHP and showed endotype-specific coupling to circulating protein levels, consistent with peripheral neutrophil activation. Abrupt increase of HR was observed once *MMP9* expression exceeded the cohort median, suggesting that elevated *MMP9* transcription identifies a subgroup of patients at highly increased risk of progression and mortality. MMP9 protein can degrade vascular basement membrane and thus facilitate migration of circulating monocytes into the site of inflammation (38). Absence of MMP9 upregulation was observed in N-fHP samples lacking neutrophils such as PBMCs, BAL, and lung tissue transcriptomes, supporting a model in which circulating neutrophils serve as the primary source of MMP9, with downstream effects on pulmonary vascular integrity and innate immune cell trafficking. Systems analysis of genes involved in vascular basement damage and neutrophil and monocyte recruitment reveal increased gene expressions in N-fHP, further supporting a role of myeloid-mediated cross-tissue pathogenesis in this endotype.

This study has several limitations. First, current HP guidelines do little to reconcile the innate and adaptive pathway signals, only suggesting that an initial innate stimulation would be expected to yield to a later adaptive immune response (37). Although our longitudinal clinical data revealed different clinical course between two endotypes, we did not have longitudinal omics data to assess molecular alterations in each endotype and determine whether individual patients can change endotype over time. Nevertheless, molecular endotyping of either heterogeneous subgroups or consecutive clinical windows will benefit precision decision-making. Second, BAL and whole blood cells derived from the same patients were not collected. Therefore, we are unable to illustrate whether L-fHP patients determined directly by whole blood transcriptomics also exhibit higher BAL lymphocytosis, which is the group expected to experience better clinical outcomes (33). Third, Our CS exposure data is retrospective and limited in small sample size, although the samples are not biased towards patients with more severe pulmonary function impairment at baseline. Overall, large cohorts of randomized prospective trials with longitudinal clinical phenotypes, paired peripheral blood and BAL samples with omics data are warranted to elucidate the endotype-specific clinical course, BAL characterization, and response to CS exposure.

## Conclusion

Molecular characterization using cross-tissue multimodal, multi-cohort omics redefine fHP as a biologically heterogeneous disease with both adaptive and innate immune axes. The two endotypes with divergent immunologic phenotypes and clinical outcomes, especially to CS response may justify a novel and more targeted therapeutic approach.

## Author contributions

YH, SFM, JSK, and IN were responsible for study conceptualization and supervision. SFM, JSK, ES, BAR, CSB, TKP, HCM, NFM, JMS, YMS, AA, SBK, MES, ERFP, MLS, AZ, NK, ALL, MVM, JMO, and FJM were responsible for patient recruitment, clinical data and biospecimen collection. YH, TV, BAR, ISC, JS, AM, and AIS participated in data analysis: YH, SFM, and IN developed the first draft of the manuscript. All authors reviewed and approved the final version of the manuscript.

## Conflicts of interest

JSK reports a grant from the National Heart, Lung, and Blood Institute, is the recipient of a Pulmonary Fibrosis Foundation Scholar’s Award, Chest Pulmonary Fibrosis Research Award; JMS reports a grant from the National Heart, Lung, and Blood Institute and Boehringer Ingelheim, is the recipient of a Pulmonary Fibrosis Foundation Scholar’s Award; AA has received research grants from the Pulmonary Fibrosis Foundation, the American College of Chest Physicians, and the National Institutes of Health for the conduct of studies in pulmonary fibrosis; serves on a pulmonary fibrosis educational forum for Boehringer Ingelheim, and has served on advisory boards to Roche, Inogen, Baxter, Brainomix, Medscape, Abbvie, patientMpower, PureTech, and Boehringer Ingelheim; MES has grants from National Institutes of Health and Boehringer Ingelheim, serves as a consultant to Boehringer Ingelheim; and participates on advisory board to Fibrogen and Bristol Mayers Squibb; MLS has grant from Boehringer Ingelheim, serves as a consultant to Boehringer Ingelheim, Trevi Therapeutical, and PureTech Health; NK reports grants from National Institutes of Health; serves as a consultant to Boehringer Ingelheim, Third Rock, Pliant, GlaxoSmithKline, Three Lake Partners, Merck, Astra Zeneca, RohBar, Biotech, Galapagos, Chiesi, Arrowhead, Sofinnova, Fibrogen, and Nuvectis, Baobab, and Brezza, reports Equity in Pliant, and grants from Astra Zeneca, Three Lakes Foundation, and BMS; JMO has grants from National Institutes of Health and Boehringer Ingelheim; serves as a consultant to Boehringer Ingelheim, Insmed, Mediar, and Avalyn; FJM reports grant support from the National Institutes of Health, other support from Boehringer Ingelheim, Biogen, Bristol-Myers Squibb, DevPro, GlaxoSmithKline, Nitto, Promedior/Roche, Vicore, Chiesi, and consulting fees from AstraZeneca, Boehringer Ingelheim, Bristol-Myers Squibb, Chiesi, Endeavor, Excalibur, GlaxoSmithKline, Lung Therapeutics/Aileron, Novartis, RS Biotherapeutics, Rejuversentx, Two XR, Hoffman Laroche unrelated to current work; IN reports having received grants from the National Heart, Lung, and Blood Institute and Veracyte; serves as a consultant to Boehringer Ingelheim, Sanofi, and UT Jenesis; YH, SFM, ES, BAR, CSB, TKP, HCM, NFM, YMS, TV, SBK, ISC, JS, AM, ERFP, AZ, ALL, MVM, and AIS declare no competing interests.

## Funding

R03AG085239 (YH); K23HL150301 and R01HL176659 (JSK); AG069264, AI112844, HL170961, AI176171, and AG090337 (JS); R01HL69166 and R01HL166290 (JMO); UG3HL145266 (FJM); UG3HL145266 and R01HL171918 (IN); R01HL132177, R01HL132287, R01HL167202, R01HL173369, and R01HL173869 (YMS); R01HL179312-01 (JMS); R01HL131565 (AM); American Lung Association (ISC); R01HL148437 and CO state grant APP-492585-CTGG1 (ERFP).

## Supporting information

https://github.com/yh9fj/Hypersensitivity-Pneumonitis/blob/main/Supplementary_Tables

## Data Availability

RNA-seq data has been deposited in NCBI GEO database under accession number GSE318816

https://github.com/yh9fj/Hypersensitivity-Pneumonitis

## Acknowledgement

We thank all participants in the Pulmonary Fibrosis Foundation (PFF) Patient Registry. We also thank investigators and other staff at participating PFF Care Centers for providing clinical data and blood samples, the PFF which established and has maintained the Patient Registry since 2016, and lastly, the many generous donors.

## Data and code availability

PFF RNA-seq raw reads data in .fastq format have been deposited in Sequence Read Archive (SRA) database. The raw counts data and meta data with clinical information have been deposited in NCBI Gene Expression Omnibus (GEO) database under Accession number GSE318816. NJH PBMC RNA-seq data has been deposited in GEO database GSE319562. Individual-level data for this study will be made available within 12 months of publication through BioLINCC (https://biolincc.nhlbi.nih.gov/home/). Investigators interested in accessing individual-level data before BioLINCC release should contact principal investigators (Imre Noth, MD and Fernando J. Martinez, MD) to discuss submitting a proposal to the PRECISIONS ancillary study committee. The codes used in the study are deposited with GitHub (https://github.com/yh9fj/Hypersensitivity-Pneumonitis/), and they are dependent on the data structure of the individual data set uploaded to GEO, SRA, and BioLINCC.

This article has an online data supplement, which is accessible from this issue’s table of content online at www.atsjournals.org.

## Notes

### Author Declarations

Study-specific protocols were approved at University of California at Davis (protocol #875917) and National Jewish Health (protocol HS-2946). The protocol for Pulmonary Fibrosis Foundation and biorepository participation was approved by institutional review boards at each participating site. All participants provided written informed consent. Any participants at both their local registry and the Pulmonary Fibrosis Foundation registry were excluded. All participants were diagnosed using established criteria and having undergone multidisciplinary review according to American Thoracic Society/European Respiratory Society criteria. University of Virginia institutional review board approved.

