## Supplementary material for "Two Blood-based Endotypes Reveal Divergent Clinical Outcomes of Fibrotic Hypersensitivity Pneumonitis": https://github.com/yh9fj/Hypersensitivity-Pneumonitis/blob/main/Supplementary_Tables

#### Methods

*Study cohorts*

Whole blood transcriptome and plasma proteome of fHP and IPF participants are obtained from Pulmonary Fibrosis Foundation (PFF) Patient Registry. University of California at Davis (UCD) cohort consisted of transcriptome of fHP patients. fHP patients National Jewish Health (NJH) provided peripheral blood mononuclear cells (PBM) transcriptome data. See detailed cross-tissue multi-omics data cohorts in Figure S1A. Study-specific protocols were approved at UCD (protocol #875917) and NJH (protocol HS-2946). The protocol for PFF and biorepository participation was approved by institutional review boards at each participating site. All participants provided written informed consent. Any participants at both their local registry and the PFF registry were excluded. All participants were diagnosed using established criteria and having undergone multidisciplinary review according to American Thoracic Society/European Respiratory Society criteria.1 Demographics and clinical traits of the three cohorts were summarized in Table 1.

*PFF cohort: Sample collection, RNA isolation, RNA-Seq library preparation and sequencing*

Venous whole blood was collected in PAXgene blood RNA tube (BD Biosciences, cat# 762165). RNA was extracted using PAXgene Blood RNA kit (Qiagen, cat#762164) following manufacturer's protocol. RNA quality and integrity were confirmed by Quant-iT RiboGreen RNA Assay Kit

(ThermoFisher, cat#R11490) and TapeStation RNA ScreenTape (Agilent, Santa Clara, CA) at Psomagen Inc., (https://www.psomagen.com). All RNA samples displayed an RNA Integrity Number (RIN)>7 were proceeded to cDNA library preparation following the manufacturer's guidelines. Total RNA in the amount of 0.1μg per sample was depleted of ribosomal RNA using the Illumina Stranded Total RNA Prep, Ligation with Ribo-Zero Plus (Illumina, cat# 470-2020-003-A). RNA was fragmented at 94°C for 2 min, followed by the directional (first strand) cDNA generation. Deoxy-UTP was incorporated in second strand synthesis to effectively quench the second strand during PCR amplification. After adenylation of the 3' end and ligation of adapters, fragments were selected and enriched with 13 cycles of PCR amplification. Libraries' sizes were checked by Agilent TapeStation 2200 or 4200 with an average size range of 330 to 445 bp. Libraries' concentrations were quantified by Picogreen method according to each protocol guidelines to create optimum cluster densities across every lane of every flow cell to achieve the higher quality of data on Illumina sequencing platforms. Clusters were generated using ExAmp chemistry on patterned flow cells and sequenced using the NovaSeq 6000S4 Reagent Kit v1.5 (300 cycles) on the NovaSeq 6000 system (Illumina, San Diego, CA), following the manufacturer's instructions. Approximately 60 million reads were generated for each cDNA library by 150bp paired end (PE) sequencing cycle.

*Bulk whole blood RNA sequencing data Processing*

Raw sequencing data in .fastq format were processed using the tool "AfterQC" for quality control (39). Around 90% sequences passed quality control score>Q30. Reads were aligned on human genome using STAR v2.7.10b (40). GenCode Human GRCh38 release-45 was used for human genome mapping and transcriptome annotation. The reads count data were normalized by by 'voom' algorithm implemented in R/Bioconductor package 'limma' to alleviate between sample variations (41). Batch effect of different RNA-seq cohort was corrected using "ComBat_seq" algorithm in R/Bioconductor package 'sva' (42).

*Bronchoalveolar Lavage (BAL) scRNA-seq and pseudo bulk RNA-seq data analysis*
R/Bioconductor package "Seurat" version-5.1.0 (43) was used to process the BAL scRNA-seq data in GSE271789 (20). One of the 10 BAL HP samples (BALHN6) have no reads count data as indicated in the published article thus was excluded from downstream data analysis. Cells annotated in original article were stratified by the fHP endotypes in current study. Reads count data retrieved from "Seurat object" of GSE271789 were aggregated into pseudo bulk RNA-seq data, and normalization by "voom" algorithm (41).

*PFF cohort: Proteomics assay and data processing*
Peripheral blood was collected in EDTA tubes. Plasma was isolated, aliquoted, and stored at $80^{O}$C. The Olink® Explore 3072 panel (Uppsala, Sweden) was used to generate semi-quantitative proteomic data for 2939 analytes covering 2921 proteins. Frozen plasma from all centers was consolidated and randomized based on center, age, sex, and race at the time of plating and processed in a single batch to mitigate batch effects. The Olink® Explore 3072 panel (Uppsala, Sweden) was used to generate semi-quantitative proteomic data for 2939 analytes covering 2921 proteins. Proteins below the lower detection limit were imputed to the lowest observed value. Protein data were normalized to minimize both intra- and inter-assay variation. Protein levels are summarized to NPX (Normalized Protein eXpression) in Log2 scale for data aggregation across plates.

*Fibrotic HP classifier construction and machine-learning (ML) prediction of endotype*

We employed the Recursive Feature Elimination (RFE) procedure in R/CRAN package 'caret' to identify the relevant classifier features (20). The 'caret' package in R facilitates a process of backward selection where less important predictors are gradually eliminated based on their importance ranking. This is determined by an external estimator four procedures: First, ranking features based on their importance scores in the model of Receiver-Operating-Characteristic curves (rocc) incorporated with repeated cross-validation (CV); Second, removing redundant features with correlation coefficient >0.7 to mitigate multi-collinearity; Third, prioritizing protein variables by the Random Forest 'rfFuncs' function in conjunction with repeated CV within the 'rfe' function in "caret" package; Fourth, retrieving key gene features and enhancing predictor selection for our analyses. Endotype classification in training cohort and prediction in test cohorts were performed using Imbalanced Random Forest (Imbalanced-RF) model implemented in R/CRAN package (44). The q-classifier assigns a sample to the minority class if the minority class conditional probability exceeds $0<q^*<1$, where $q^*$ equals the unconditional probability of observing a minority class sample (45).

*Immune cells deconvolution*

We used R package 'immunedeconv' to quantify the fraction the immune cell types from bulk RNA-sequencing data (21). Transcript per Million (TPM) data normalizing both sequencing depth and gene length were utilized for immune cells deconvolution. Specifically, we used the 'quanTIseq' method, which has been extensively validated in blood samples using simulated, flow cytometry, and immunohistochemistry data (46).

*Survival analysis of endotypes and corticosteroid exposure*

Kaplan-Meier (KM) analyses were used for unadjusted comparisons of TFS between each fHP endotype and idiopathic pulmonary fibrosis (IPF). Hazard ratios (HR) and 95% confidence intervals (CI) were estimated using Cox proportional hazards (Cox-PH) regression models.

Univariate Cox-PH models were used for pairwise survival comparisons, while multivariable Cox-PH models were used to adjust for potential confounders.

Corticosteroid (CS) exposure was defined as systemic CS treatment initiated prior to blood sampling. The association between CS exposure and TFS was evaluated separately within each fHP endotype using KM analysis with log-rank testing and Cox-PH regression. To assess whether the effect of CS exposure on TFS differed by fHP endotype, an interaction term between CS exposure and endotype was included in a multivariable Cox-PH model adjusting for age, sex, FVC-pp, DLCO-pp. Endotype-stratified CS effect was computed using delta method implemented in R package function multcomp::glht() (47).

*Differential expression analysis*

Differential expression of genes or proteins between two groups were identified using empirical Bayesian-moderated t-test implemented in R/Bioconductor package “limma” (48-49). P-values were adjusted for multiple testing using Benjamini-Hochberg method (50). The criterion of differentially expressed genes was set at false discover rate (FDR) <0.05.

*Functional pathway analysis*

Gene set enrichment analysis (GSEA) Gene set enrichment analysis (GSEA) between two groups was conducted using R/Bioconductor package “clusterProfiler” version-4.0.5 (51-52). Gene Ontology biological processes, KEGG and Wiki pathways databases were used to determine whether an a priori defined gene set demonstrated statistically significant differences between two biological states.

*Principal Component Analysis (PCA) and projection*

PCA was performed using R CRAN package “FactoMineR” with the lowest p-value threshold at $p=2.2x10^{-16}$. PCA projection was performed by computing principal components and projecting both reference and query datasets onto these components to visualize their similarity in reduced-dimensional space. To account for sampling variability and improve robustness of the projections, bootstrap resampling with 100 iterations was performed prior to PCA computation.

*Other Statistics*

Data were reported as Mean±SD (standard deviation) for continuous variables and frequencies (percentages) for categorical variables. T-test or Wilcoxon-test was used to analyze continuous variables, depending on data distribution. Chi-squared or Fisher's exact test was used for categorical data, as appropriate.

#### RESULTS

*Functional characterization of the fHP endotypes reveals distinct adaptive versus innate immune programs*

Wiki pathways showed that Minor (N-fHP) group was associated with innate immune functions such as IL1 signaling, while Major (L-fHP) group involved adaptive immune responses such as activation of T cells receptor signaling and Th17 cell differentiation in both PFF and UCD WB transcriptome cohorts (Figure S3A-B). PBMCs transcriptome of the NJH cohort, which lacks neutrophils, exhibited lymphocyte mediated immunity in Major (L-fHP), and cilium assembly in Minor (N-fHP) (Figure S3C).

We further compared lung biopsies gene expression patterns (GSE150910; Figure S1A) between the two groups classified in Figure 1H and identified 5463 DEG including 2735 up-regulated and 2728 down-regulated genes in Minor (N-fHP) compared to Major (L-fHP) (Figure S4A, Table S5). GSEA of Gene Ontology biological process confirmed adaptive immune response and lymphocyte activation in L-fHP,, and further revealed collagen-containing extracellular matrix (ECM) activities in N-fHP endotype (Figure S4B).

*Transcriptomic fHP-endotypes in the PFF cohort differ in baseline disease severity, longitudinal outcomes and corticosteroid (CS) response*

Unsupervised PCA further confirmed that the fHP endotype classification was not contingent upon corticosteroid usage prior to blood draw (Figure S7A-C), or UIP status (Figure S7D). Findings suggested that these clinical traits were not determinant of endotype.

N-fHP patients also exhibited significantly lower baseline FVC-pp in the NJH cohort (AUC=0.81, p=0.028, Figure S6A&C). A similar trend was observed in the UCD cohort, although it did not reach statistical significance (AUC=0.62, p=0.22, Figure S6B&C). Kaplan-Meier (KM) analyses of TFS showed no significant difference between the two endotypes in UCD (log-rank p=0.67, Figure 6D). The lack of statistical significance in the UCD cohort likely reflects limited follow-up duration (mean <12 months), small sample size (N=40) and events numbers (N=13), resulting in insufficient power to detect the true differences (Table 1).

*BAL single-cell transcriptomics recapitulates blood transcriptome-defined fHP endotypes*

GSEA of the endotypes in BAL pseudo-bulk RNA-seq data affirmed the two immune subtypes similar as observed in the WB transcriptome cohorts. GSEA of GO Bioprocess confirmed T cell differentiation, lymphocyte differentiation, and T cell activation in L-fHP, while neutrophil and myeloid cell activation and neutrophil mediated immunity are shown in N-fHP (Figure S9A). KEGG pathways analyses confirmed activated T cell receptor signaling pathway in L-fHP, and cytokine-cytokine receptor interaction in N-fHP (Figure S9B). Thus, the pathway analyses validated endotypes in the BAL scRNA-seq data.

**Table S1. fHP-endotype classifier derived from the transcriptome of PFF cohort**

| Symbol | %BS# | $Log_2FC$ | AvgExpr | p.value | adj.p.value |
|---|---|---|---|---|---|
| *TENM1* | 89 | 1.63 | 3.35 | 6.43E-15 | 4.98E-14 |
| *PRUNE2* | 95 | 1.59 | 0.78 | 1.34E-15 | 1.15E-14 |
| *ARMC12* | 87 | 1.51 | 1.29 | 3.34E-17 | 3.85E-16 |
| *S100P* | 95 | 1.38 | 6.24 | 1.37E-08 | 4.25E-08 |
| *C5orf47* | 86 | 1.18 | 0.03 | 4.24E-16 | 4.01E-15 |
| *POU5F1* | 89 | 1.08 | 1.13 | 9.39E-14 | 6.01E-13 |
| *SLC22A14* | 94 | 0.92 | 0.30 | 7.30E-15 | 5.60E-14 |
| *MFSD9* | 88 | 0.90 | 4.52 | 1.80E-13 | 1.11E-12 |
| *TSNAX-DISC1* | 94 | 0.85 | 0.87 | 1.83E-15 | 1.54E-14 |
| *SEMA3C* | 96 | 0.78 | 3.41 | 4.09E-14 | 2.77E-13 |
| *CNIH2* | 89 | 0.77 | 1.74 | 6.48E-12 | 3.20E-11 |
| *ZNF763* | 94 | 0.74 | 3.18 | 4.89E-13 | 2.82E-12 |
| *CNIH3* | 86 | 0.66 | 1.74 | 1.37E-15 | 1.18E-14 |
| *ABHD11* | 89 | 0.60 | 4.26 | 2.60E-14 | 1.81E-13 |
| *RDH5* | 89 | 0.59 | 3.32 | 7.66E-14 | 4.98E-13 |
| *CORO2A* | 85 | 0.55 | 4.16 | 2.76E-14 | 1.92E-13 |
| *BLOC1S1* | 86 | 0.54 | 4.93 | 6.02E-16 | 5.54E-15 |
| *COA1* | 99 | 0.52 | 5.07 | 1.86E-16 | 1.86E-15 |
| *ZSCAN16* | 88 | 0.46 | 4.85 | 1.77E-13 | 1.09E-12 |
| *AKIRIN2* | 95 | 0.24 | 7.93 | 3.05E-11 | 1.35E-10 |
| *ZSCAN32* | 91 | 0.24 | 4.81 | 1.82E-13 | 1.12E-12 |
| *DNAJC2* | 89 | -0.28 | 3.29 | 1.45E-10 | 5.81E-10 |
| *SEH1L* | 86 | -0.30 | 3.36 | 1.82E-15 | 1.53E-14 |
| *EI24* | 88 | -0.30 | 4.68 | 3.91E-14 | 2.65E-13 |
| *ACTR5* | 92 | -0.43 | 4.23 | 8.40E-15 | 6.36E-14 |
| *MRPS2* | 89 | -0.43 | 4.36 | 1.38E-16 | 1.42E-15 |
| *SMDT1* | 89 | -0.46 | 5.50 | 2.96E-13 | 1.76E-12 |
| *RHNO1* | 89 | -0.46 | 4.26 | 1.20E-14 | 8.88E-14 |
| *SLC35F2* | 85 | -0.66 | 2.47 | 4.73E-14 | 3.17E-13 |
| *RRP7A* | 97 | -0.69 | 5.80 | 2.20E-14 | 1.56E-13 |
| *PAFAH1B3* | 93 | -0.69 | 3.94 | 3.91E-16 | 3.72E-15 |
| *SALL2* | 86 | -0.71 | 1.43 | 2.60E-10 | 1.01E-09 |
| *SLC4A3* | 85 | -0.73 | 0.33 | 1.54E-11 | 7.18E-11 |
| *BYSL* | 98 | -0.75 | 3.08 | 1.49E-16 | 1.51E-15 |
| *SGPP2* | 94 | -0.79 | 2.56 | 3.97E-14 | 2.69E-13 |
| *AMZ1* | 93 | -0.87 | 0.98 | 4.33E-13 | 2.51E-12 |
| *CEP170B* | 86 | -0.88 | 1.11 | 1.24E-14 | 9.15E-14 |
| *RNF207* | 96 | -1.04 | 1.46 | 1.44E-17 | 1.77E-16 |
| *SPON1* | 93 | -1.05 | 0.76 | 7.53E-16 | 6.82E-15 |
| *IGFBP4* | 96 | -1.21 | 2.52 | 3.30E-14 | 2.26E-13 |

### BS: Bootstrap sampling of equal number of N-fHP and L-fHP cases for Recursive Feature Elimination.
### %BS: represents the percentage of inclusion in 100 bootstrap sampling of RFE for classifier construction. Genes colored in red and blue represent up- and down-regulated genes in N-fHP compared to L-fHP, respectively.

**Table S2. Validation of fHP 40-gene classifier by Imbalanced Random Forrest**

| Cohort | Prediction | N-fHP | L-fHP | Sensitivity (%) | Specificity (%) |
|---|---|---|---|---|---|
| **PFF** 5-fold cross-validation | Predicted N-fHP | 37 | 3 | 97.4 | 96.6 |
| | Predicted L-fHP | 1 | 85 | | |
| **UCD** Independent validation | Predicted N-fHP | 10 | 1 | 66.7 | 96.0 |
| | Predicted L-fHP | 5 | 24 | | |

**Supplementary Table S3-S7 download –**

https://github.com/yh9fj/Hypersensitivity-Pneumonitis/tree/main/Tables_S3-7

**Figure S1**

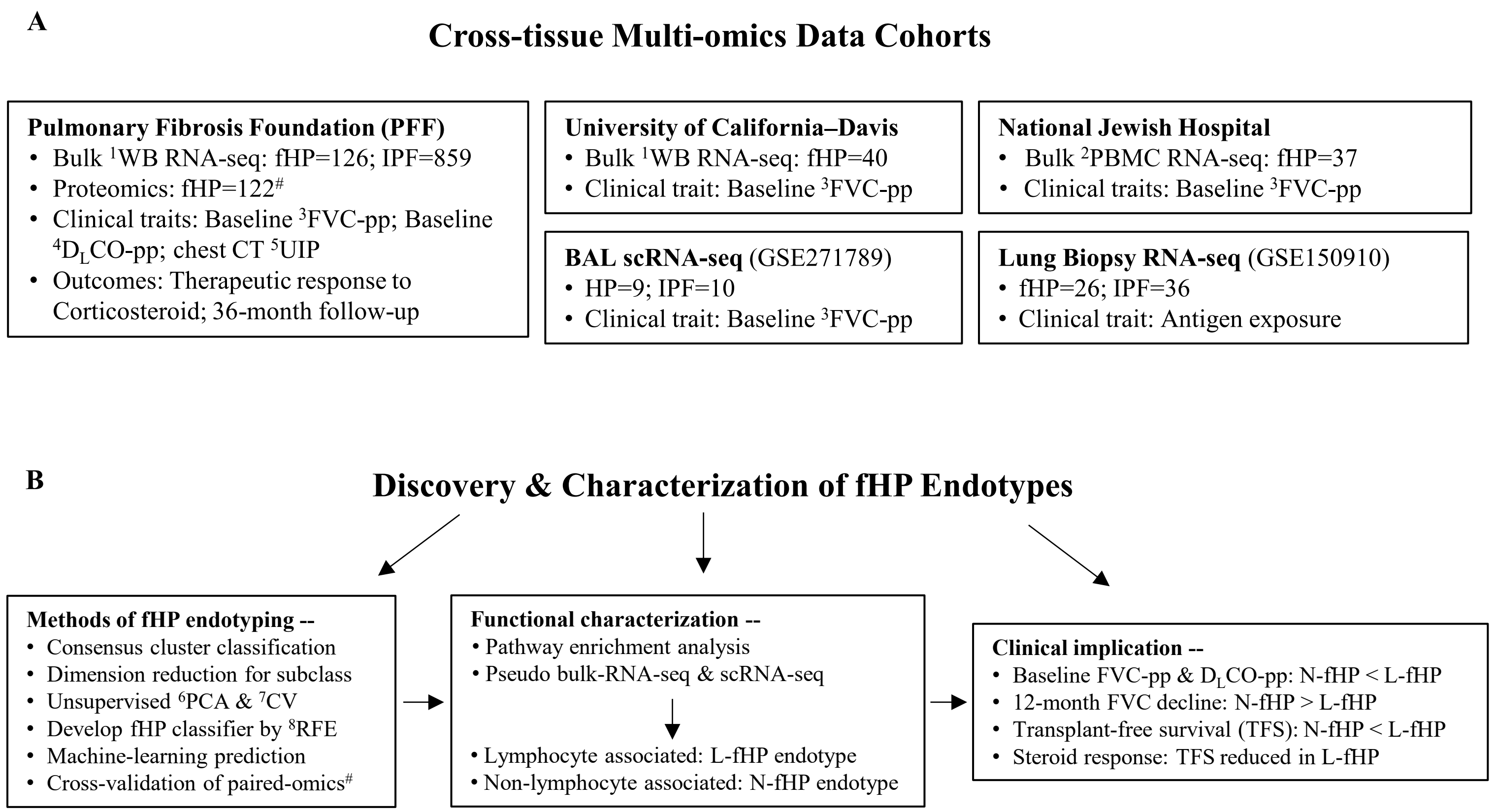


[1]WB=Whole Blood
[2]PBMC=Peripheral Blood Mononuclear Cells
[3]FVC-pp=Forced Vital Capacity Percent Predicted
[4]$D_L$CO-pp=Diffusing capacity of the Lungs for Carbon Monoxide Percent Predicted
[5]UIP=Usual Interstitial Pneumonia
[6]PCA=Principal Component Analysis
[7]CV=Cross-Validation
[8]RFE=Recursive Feature Elimination with bootstrap sampling procedure to prioritize genes for machine learning model
[#] Proteomic data and bulk whole blood RNA-seq data were derived from the same patients in PFF cohort

**Figure S1. Study design.** (A) Summary of the multi-omics cohorts with clinical data across multiple tissues. Paired transcriptomic and proteomic data were generated from the Pulmonary Fibrosis Foundation (PFF) Patient Registry. Transcriptomic data from University of California-Davis (UCD), National Jewish Hospital (NJH), Bronchoalveolar lavage (BAL), and surgical lung biopsies were used as validation cohorts. (B) Methods of fHP endotyping, identification, validation, functional characterization, pathway enrichment analysis, cell type deconvolution, and clinical association and implication of the novel fHP endotypes.

**Supplementary Figure S2**

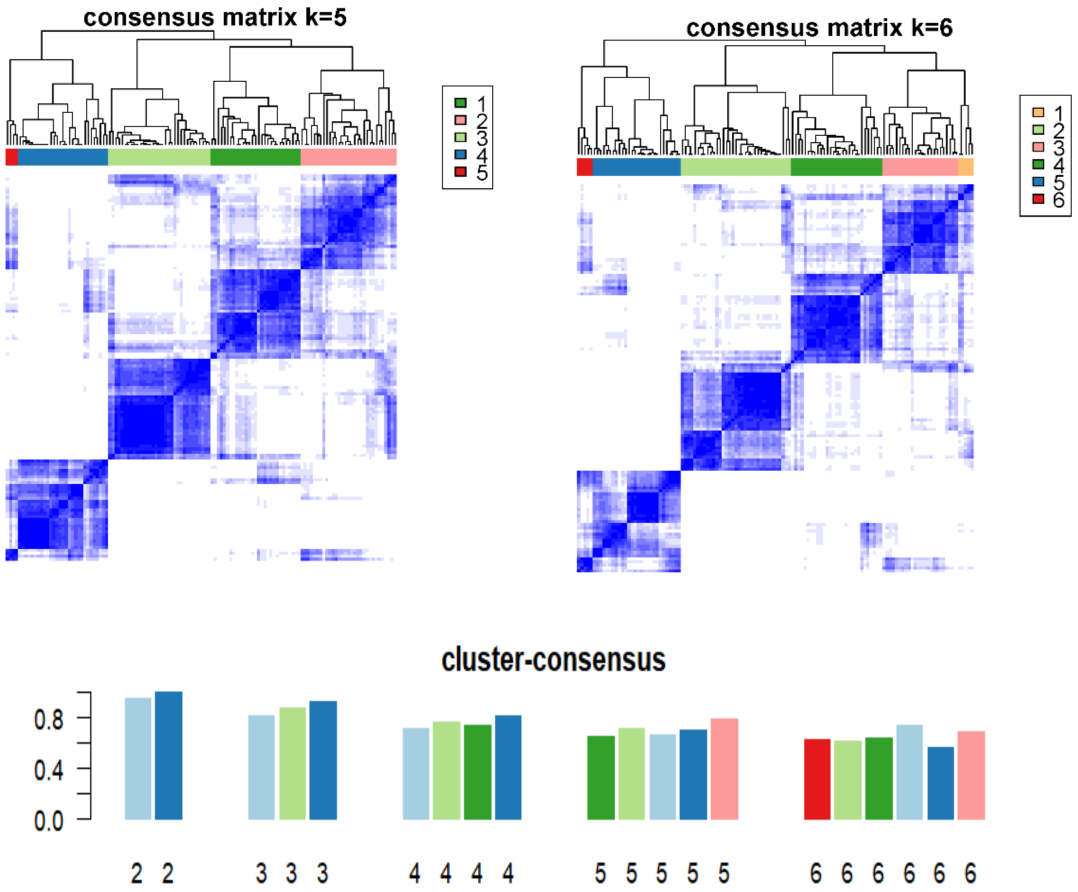


| | DDRTree+ | DDRTree- | Sensitivity | Specificity |
|---|---|---|---|---|
| Consensus clustering+ | 33 | 0 | 86.8% | 100% |
| Consensus clustering- | 5 | 88 | | |

**Figure S2. Determination of optimal number of clusters in PFF cohort using unsupervised machine learning Consensus clusters.** (A-B) Clustering distribution of 100 subsampling iteration with preset cluster number of k=5-6. See Figure 1A-C for k=2-4. (C) Bar plot displays the mean of within-cluster consensus scores for clustering stability, with cluster k=2 showing the highest stability score. (D) Cross-validation between DDRTree with Consensus clustering k=2.

**Figure S3**

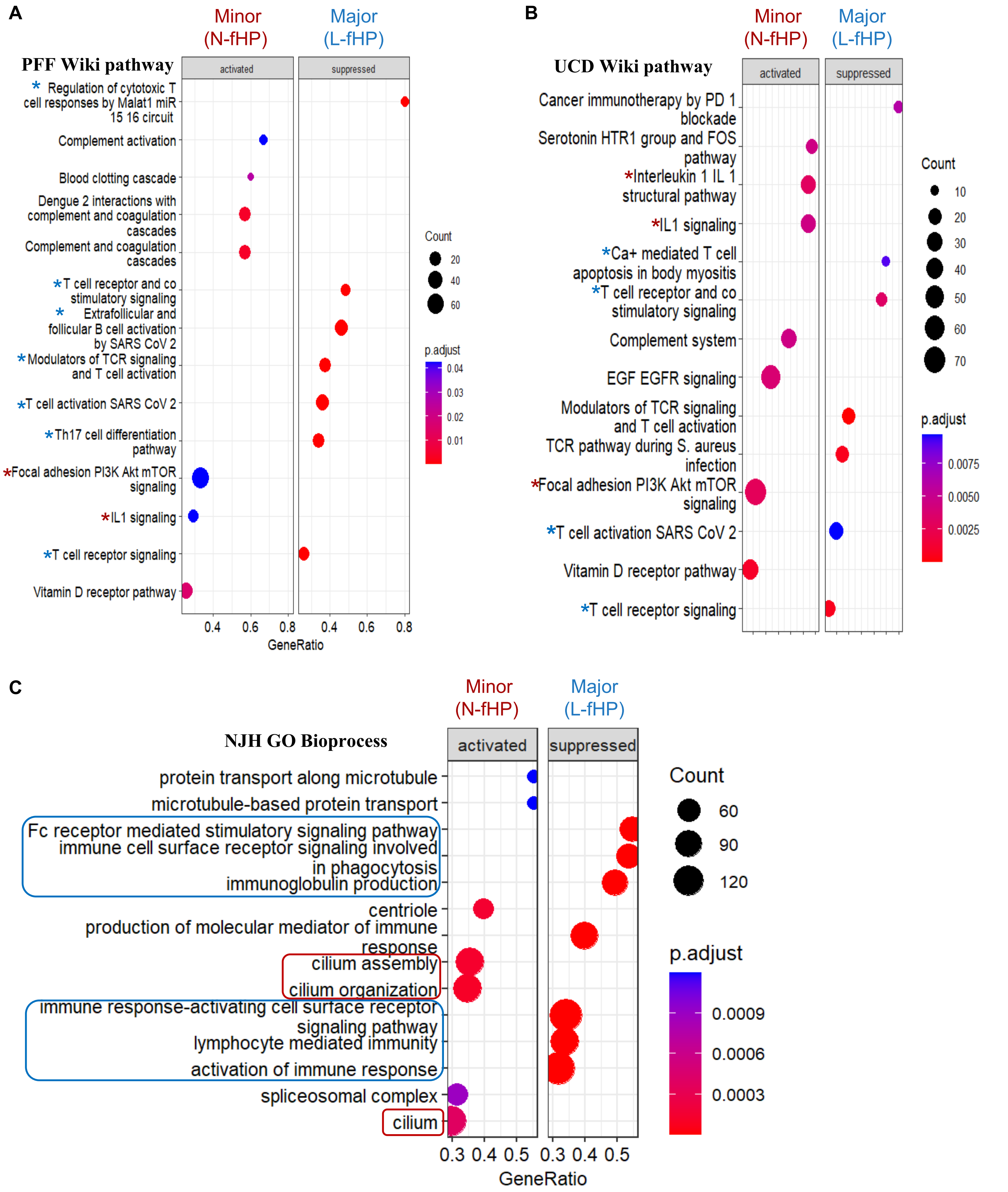


**Figure S3. Gene Set Enrichment Analysis (GSEA) of PFF and UCD transcriptome.** Wiki pathway analysis in **(A).** PFF and **(B).** UCD cohort with bulk whole blood RNA-seq confirmed neutrophil mediated immunity and IL1 signaling pathway activation in Minor/N-fHP (red asterisk)**,** and T cell and B cell activation in Major/L-fHP (blue asterisk). C. Gene Ontology analysis of NJH cohort with bulk PBMCs RNA-seq demonstrated lymphocyte mediated immunity in Major/L-fHP (blue box), and cilium activities in Minor/N-fHP (red box).

**Figure S4**

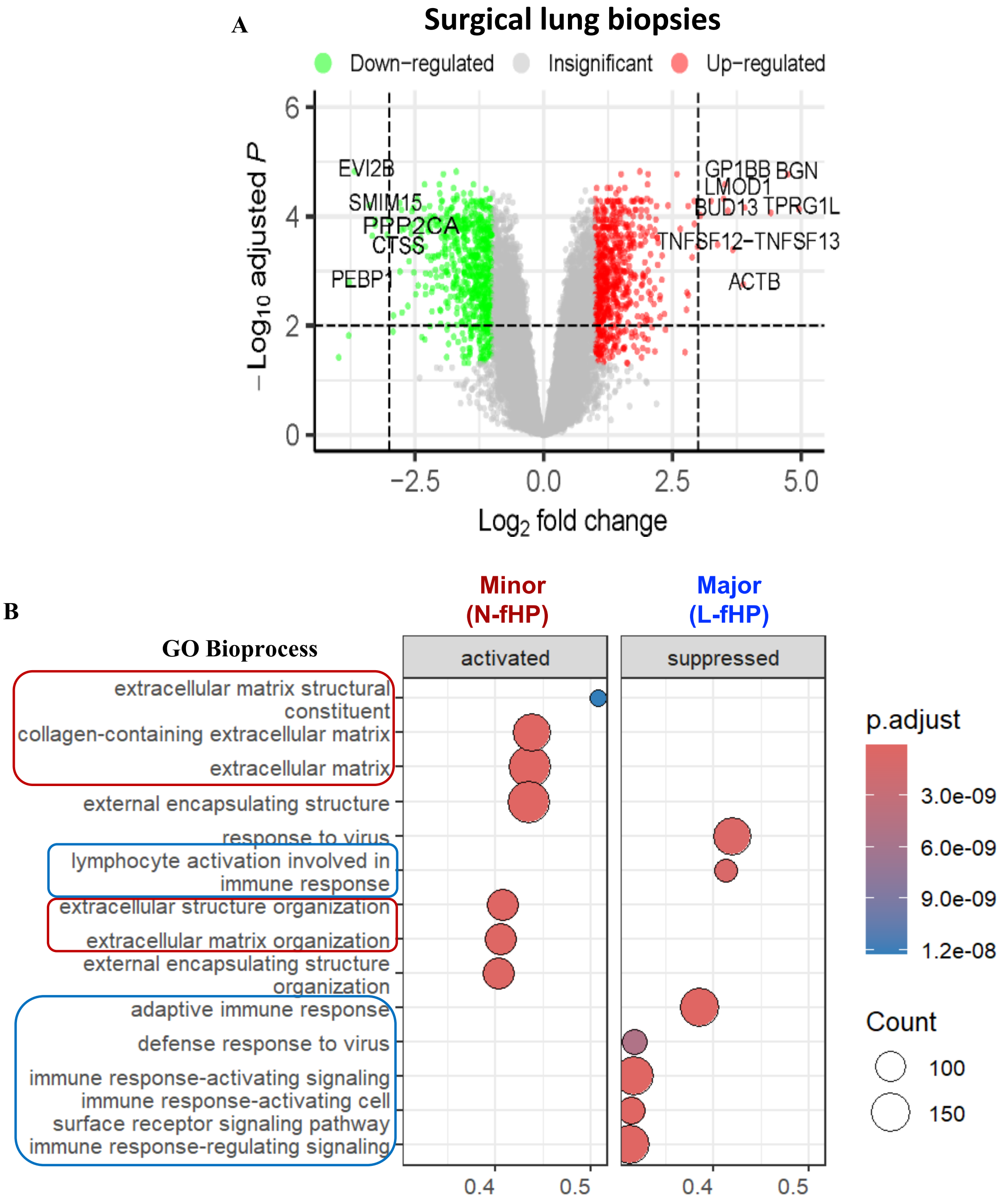


**Figure S4 Volcano plot and Gene Set Enrichment Analysis (GSEA) between fHP endotypes of RNA-seq from surgical lung biopsies (GSE150910).** A. Volcano plot of DEG identified by empirical Bayes moderated t-test between two endotypes in lung biopsies. Red and green colors represent up- and down-regulated genes with fold change (FC) > 2 and false discovery rate (FDR) < 0.05 in Minor compared to Major, respectively (See Fig. 1i). The full list of 5463 with FDR < 0.05 is displayed in Supplementary Table 5. **B.** GSEA for GO biological process confirmed lymphocyte activation pathway, adaptive immune response and Th17 cell differentiation in Major (L-fHP) endotype of lung biopsies (blue box). Collagen-containing extra-cellular matrix (ECM) activity and ECM-receptor interaction were increased in N-fHP endotype of lung biopsies (red box).

**Figure S5**

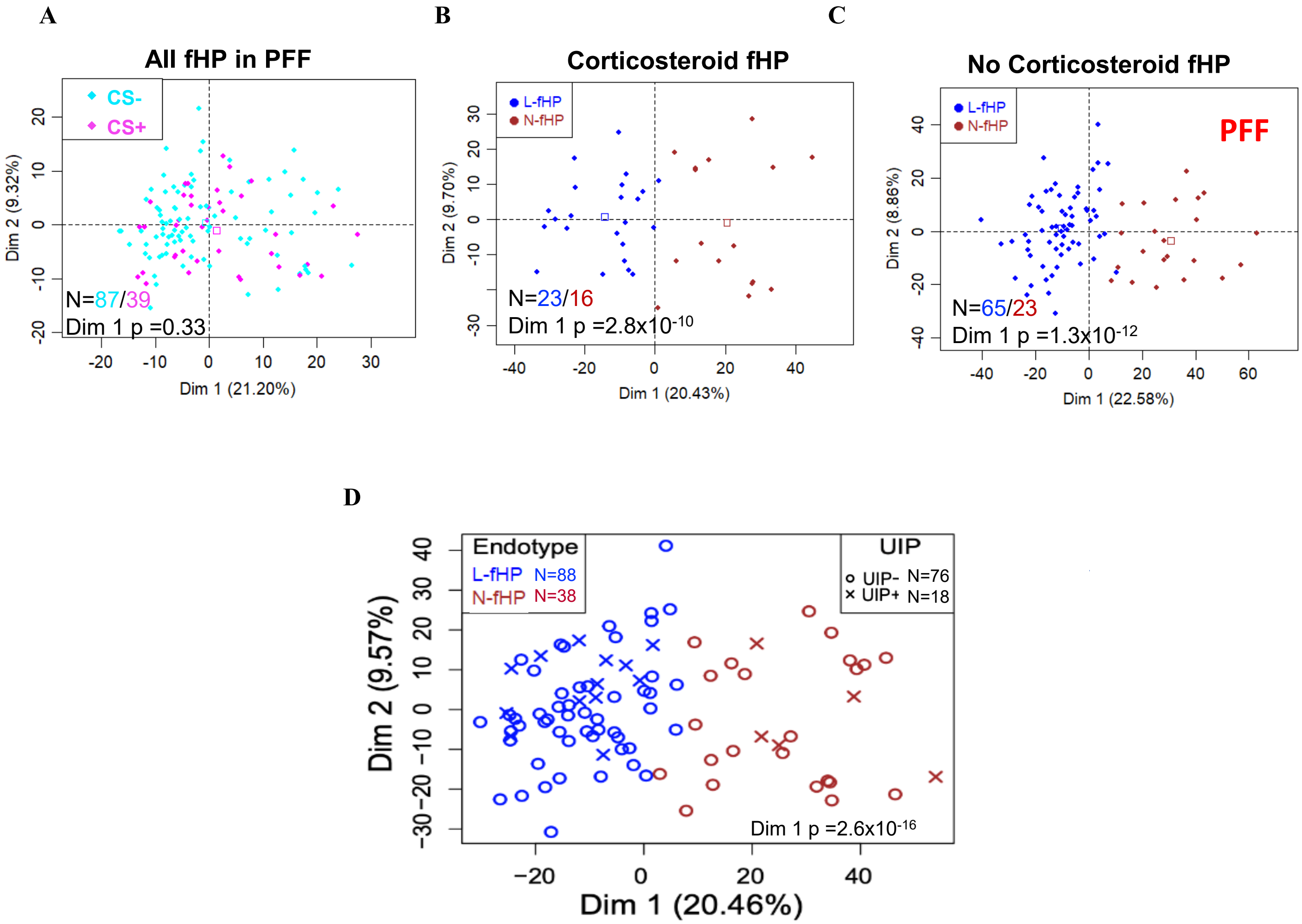


**Figure S5. Unsupervised Principal Component Analysis (PCA) of fHP patients in PFF cohort.** (A) All fHP patients in PFF cohort stratified by corticosteroid (CS) exposure prior to blood draw exhibited no significant difference (p=0.33). (B) Endotype-stratified fHP patients exposed to CS prior to sample draw. (C) Endotype-stratified fHP patients not exposed to CS. (D) fHP patients stratified by endotype (red or blue color) and CT UIP status (-/+) denoted by circle or cross, respectively**.** Case numbers of L-fHP/N-fHP and *t*-test p-value of Principal Component-1 (Dim 1) are displayed in graph B-D. See Table S5 for detail.

**Figure S6**

**A**

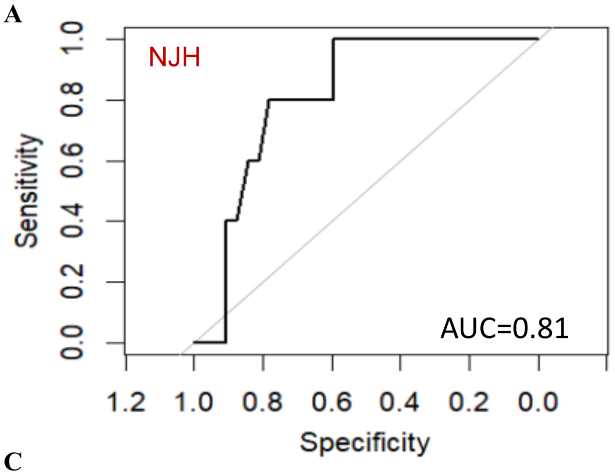


**B**

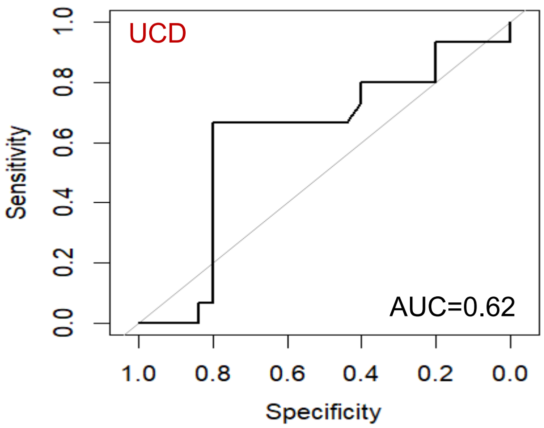


**C**

| FVC-pp | NJH | UCD |
|---|---|---|
| L- fHP (Mean±SD) | 61.2±10.8 | 63.2±21.8 |
| N- fHP (Mean±SD) | 74.2±12.6 | 58.5±18.5 |
| T-test p-value | 0.028 | 0.22 |

**D**

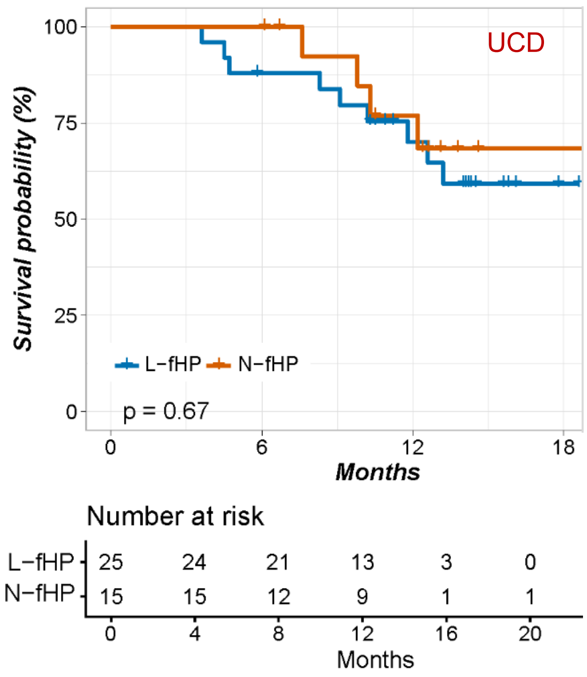


**Figure S6. Clinical association of endotypes in external cohorts.** Receiver Operating Characteristic (ROC) analysis in (A) NJH cohort and (B) UCD cohort. (C) Baseline FVC-pp between L-fHP and N-fHP endotypes demonstrating a significant difference in the NJH cohort (p=0.028), but not in the UCD cohort (p=0.22). (D) Kaplan-Meier (KM) analysis of transplant-free survival (TFS) between L-fHP and N-fHP endotypes in UCD cohort (log-rank p=0.67).

**Figure S7**

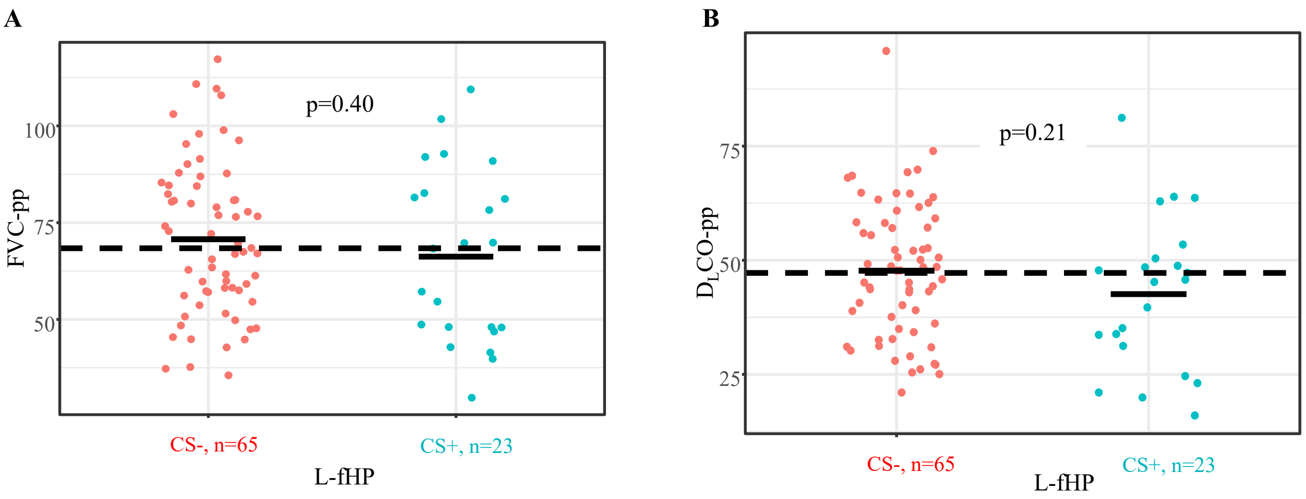


**Figure S7. Baseline pulmonary function stratified by corticosteroid exposure in L-fHP endotype of the PFF cohort.** (A) Baseline FVC-pp or (B) $D_L$CO-pp did not differ in L-fHP endotype with or without CS exposure initiated before blood sampling.

**Figure S8**

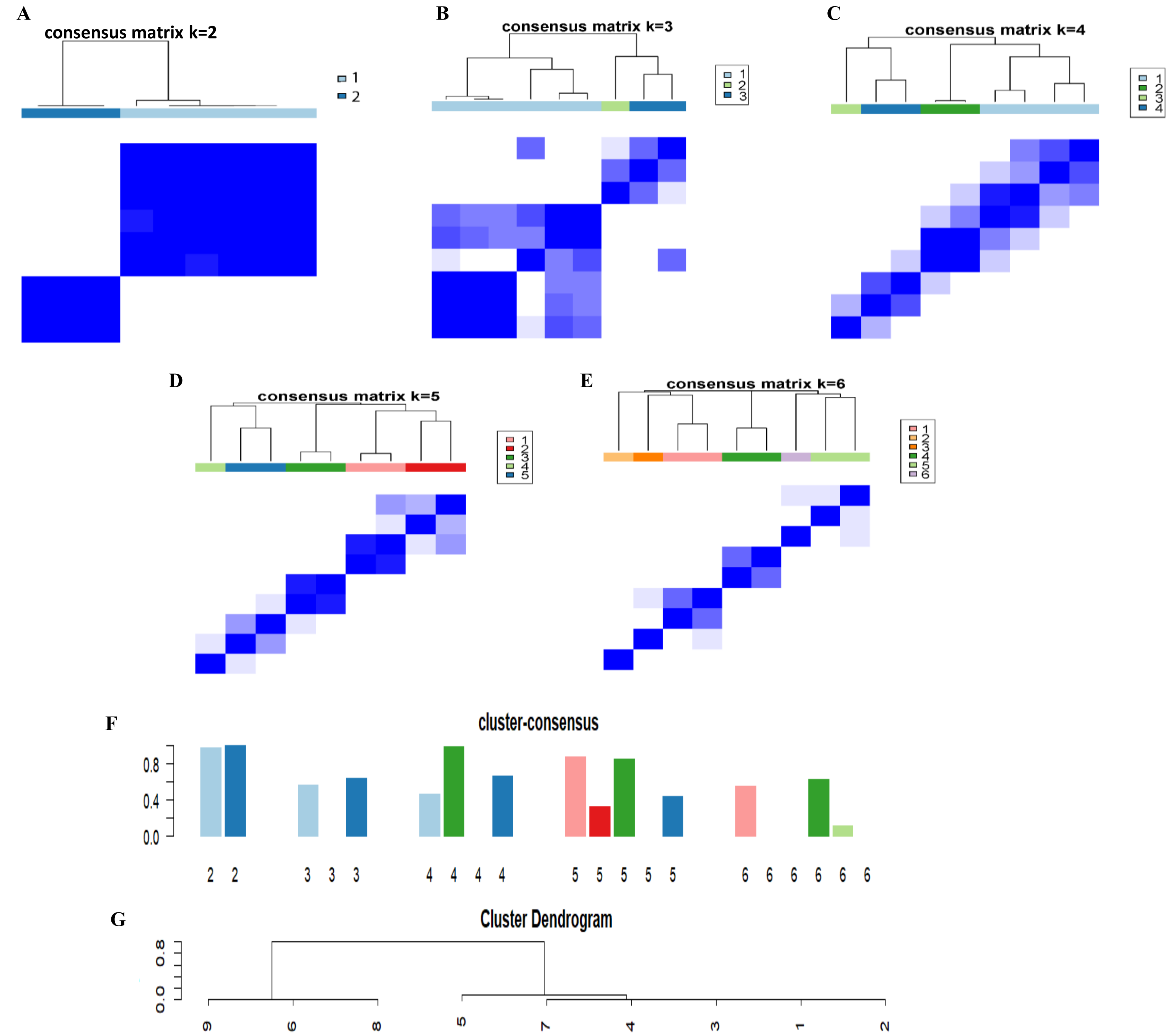


**H**

| | DDRTree+ | DDRTree- | Sensitivity | Specificity |
|---|---|---|---|---|
| Consensus clustering+ | 6 | 0 | 100% | 100% |
| Consensus clustering- | 0 | 3 | | |

**Figure S8. Determination of optimal number of clusters in BAL samples (GSE271789) using unsupervised machine learning (ML) Consensus clusters.** Pseudo-bulk RNA-seq data were generated from scRNA-seq of GSE271789 BAL samples. Clustering distribution of 100 subsampling iteration with preset cluster of k=2-6. (A) Consensus cluster k=2 displayed a clearer border, indicating most stable classification. (B) Consensus cluster k=3. (C) Consensus cluster k=4. (D) Consensus cluster k=5. (E) Consensus cluster k=6. (F) Bar plot displays the mean of within-cluster consensus scores for clustering stability, with cluster k=2 showing the highest stability score. (G) Hierarchical clustering of the BAL samples based on ML consensus. (H) Cross-validation between DDRTree with Consensus clustering when k=2.

**Figure S9**

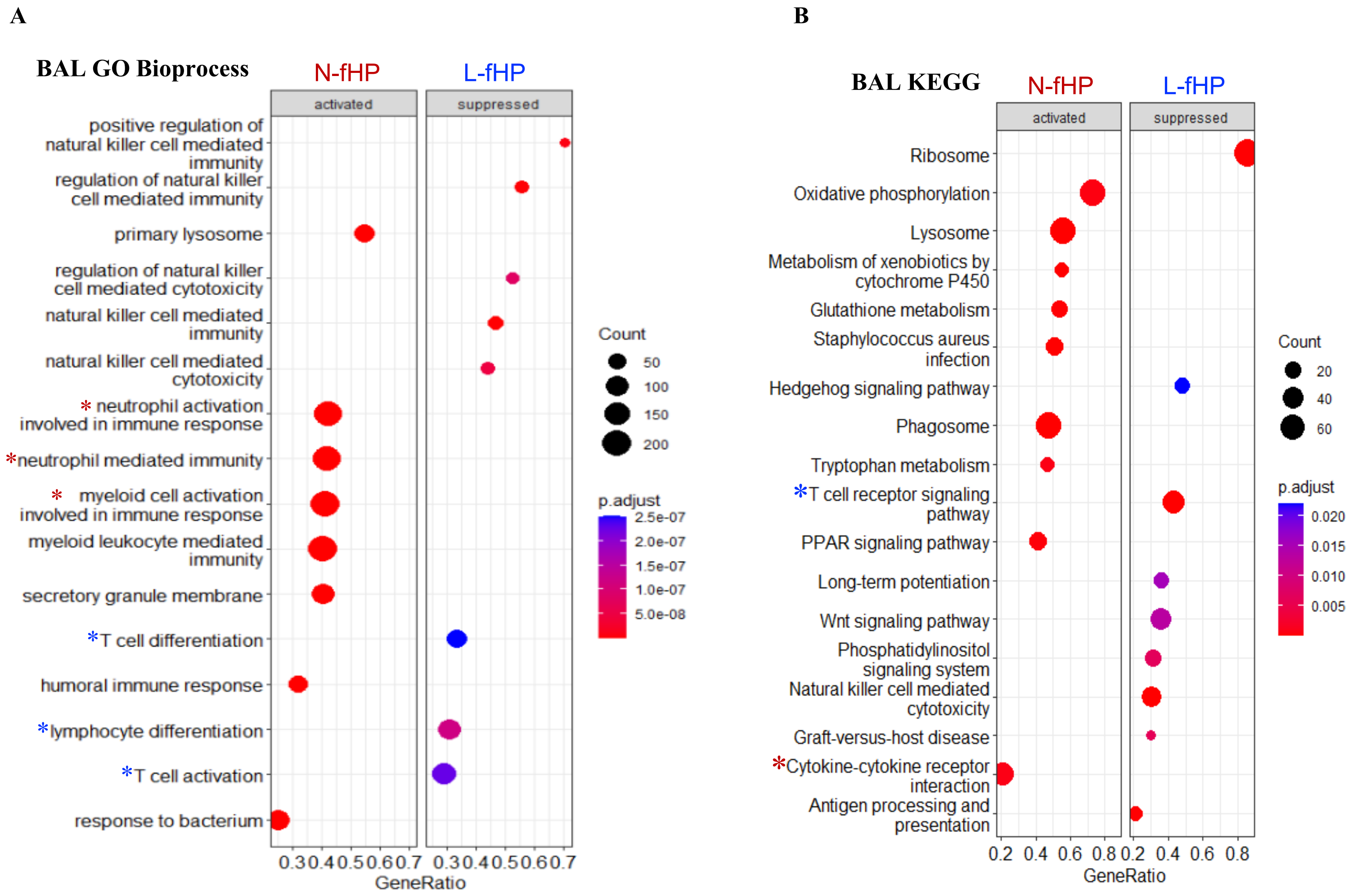


**Figure S9. Gene Set Enrichment Analysis (GSEA) between fHP endotypes in Bronchoalveolar leverage (BAL) transcriptomic samples.** (A) GSEA of Gene Ontology (GO) Bioprocess confirmed T cell differentiation, lymphocyte differentiation, and T cell activation in L-fHP (blue asterisks); neutrophil and myeloid cell activation and neutrophil mediated immunity in N-fHP (red asterisks). B. KEGG pathways confirmed activated T cell receptor signaling pathway in L-fHP (blue asterisks), and cytokine-cytokines receptor interaction in N-fHP (red asterisks).

**Figure S10**

**A**

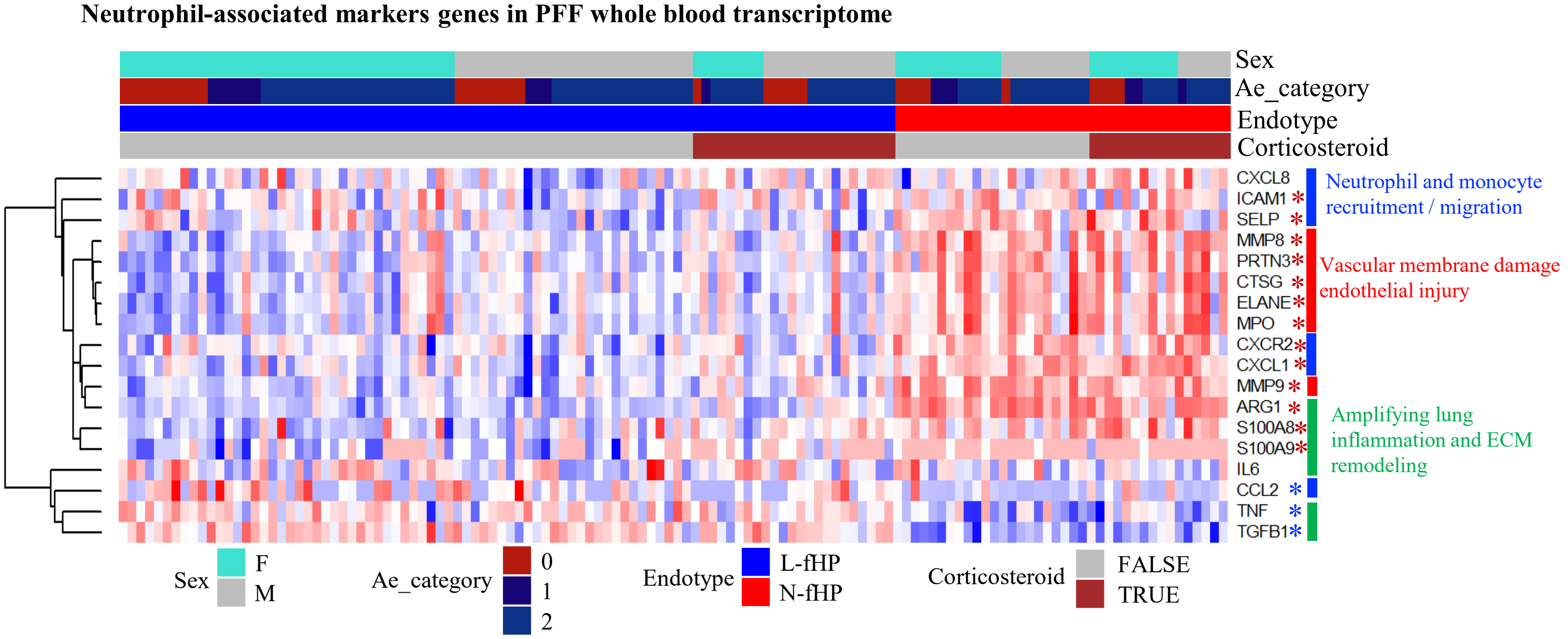


**B**

| Symbol | Gene name | Biological Function | PMID-1 | PMID-2 |
|---|---|---|---|---|
| ELANE | Elastase, neutrophil expressed | Vascular membrane damage; endothelial injury | 25743626 | 22441751 |
| CTSG | Cathepsin G | Vascular membrane damage; endothelial injury | 41394824 | 21079042 |
| PRTN3 | Proteinase 3 | Vascular membrane damage; endothelial injury | 41394824 | 21079042 |
| MMP8 | Matrix metallopeptidase 8 | Vascular membrane damage; endothelial injury | 23487425 | 39603661 |
| MPO | Myeloperoxidase | Vascular membrane damage; endothelial injury | 34207641 | 15182268 |
| MMP9 | Matrix metallopeptidase 9 | Vascular membrane damage; endothelial injury | 35804363 | 10956632 |
| CXCL8 | C-X-C motif chemokine ligand 8 (interleukin 8) | Neutrophil and monocyte recruitment / migration | 9616535 | 9517588 |
| CXCL1 | C-X-C motif chemokine ligand 1 | Neutrophil and monocyte recruitment / migration | 26138727 | 40047857 |
| CXCR2 | C-X-C motif chemokine receptor 2 | Neutrophil and monocyte recruitment / migration | 41564599 | 40047857 |
| CCL2 | C-C motif chemokine ligand 2 | Neutrophil and monocyte recruitment / migration | 31922885 | 39955168 |
| ICAM1 | Intercellular adhesion molecule 1 | Neutrophil and monocyte recruitment / migration | 10836326 | 34484188 |
| SELP | Selectin P | Neutrophil and monocyte recruitment / migration | 16936251 | 9777942 |
| TNF | Tumor necrosis factor | Amplifying lung inflammation and ECM remodeling | 9839161 | 12816730 |
| IL6 | Interleukin 6 | Amplifying lung inflammation and ECM remodeling | 25172494 | 35857823 |
| S100A8 | S100 calcium binding protein A8 | Amplifying lung inflammation and ECM remodeling | 34154976 | 33169236 |
| S100A9 | S100 calcium binding protein A9 | Amplifying lung inflammation and ECM remodeling | 22209187 | 33169236 |
| TGFB1 | Transforming growth factor beta 1 | Amplifying lung inflammation and ECM remodeling | 34013369 | 28432134 |
| ARG1 | Arginase 1 | Amplifying lung inflammation and ECM remodeling | 40875483 | 23637937 |

**Figure S10. Hierarchical clustering of neutrophil-associated markers in PFF transcriptome** (A) Genes involved in vascular basement damage and endothelial injury, neutrophil and monocyte recruitment or migration were predominantly expressed in N-fHP on left sample cluster. Red or blue Asterix indicates genes significantly (FDR<0.05) increased or decreased in N-fHP compared to L-fHP, respectively (see Table S3 for DEG list in PFF transcriptome). (B) Function and citation of neutrophil-associated markers in PFF transcriptome

**Figure S11**

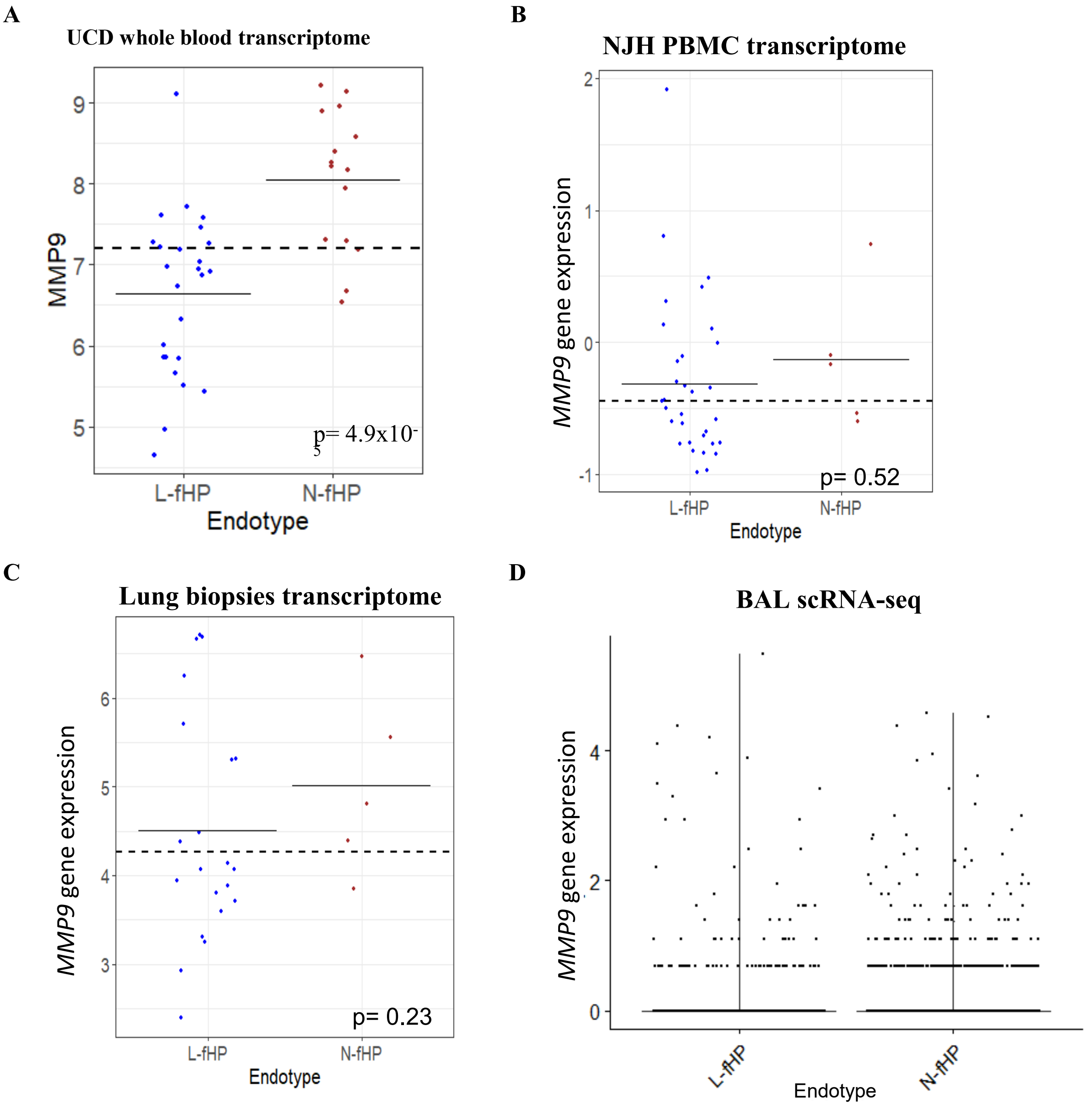


**Figure S11. Myeloperoxidase (MMP9) gene expression in diverse tissue samples.** MMP9 transcript levels in (A) UCD whole blood exhibited significantly higher transcript levels in N-fHP than L-fHP. (B) NJH PBMC,;(C) lung biopsies RNA-seq data (GSE150910), and (D) BAL scRNA-seq data that were devoid of neutrophils displayed no difference between L-fHP and N-fHP endothypes.

**Figure S12**

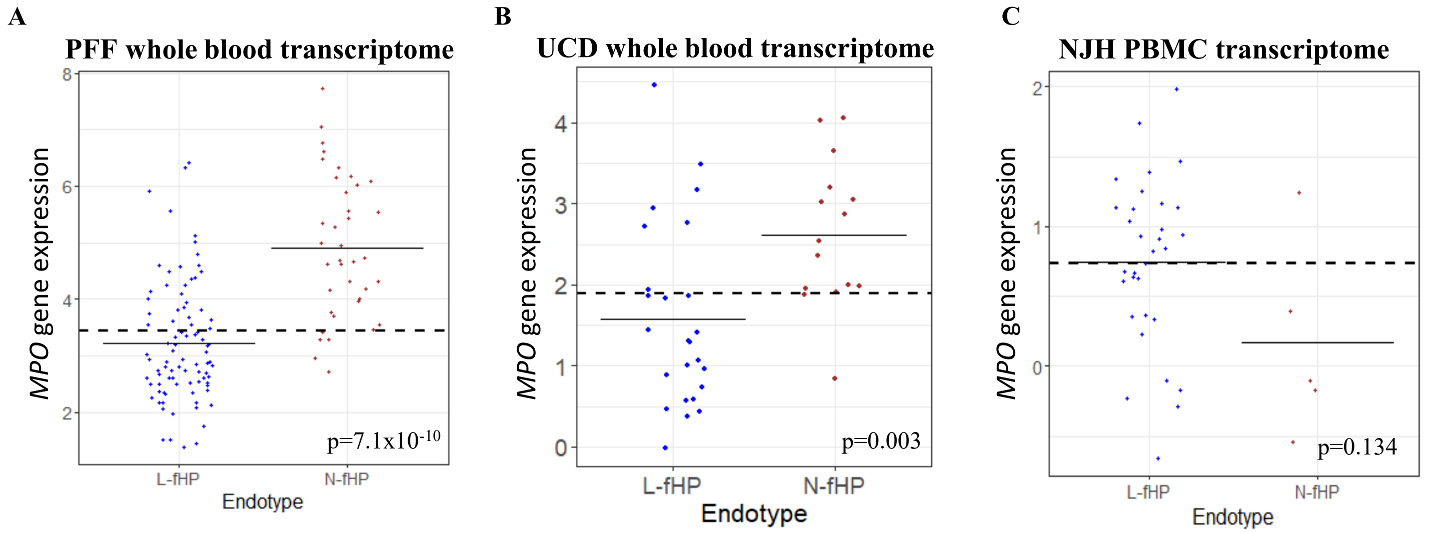


**Figure S12. Myeloperoxidase (MPO) gene expression in peripheral blood transcriptome.** MPO gene expressions displayed significant enhancement in N-fHP compared to L-fHP in (A) PFF whole blood transcriptome cohort and (B) UCD whole blood transcriptome cohort, but not in (C) NJH PBMC transcriptome cohort. MPO expressions in 87% N-fHP patients in both PFF (33/38) and UCD (13/15) were above the median levels of the corresponding cohort (A-B).
